# Copy Number Variants and their Implications for Developmental and Behavioural Problems in Cleft Lip and/or Palate

**DOI:** 10.1101/2025.04.30.25326724

**Authors:** Alexandros Rammos, Rachel Blakey, Charlotte A Dennison, Sarah J Lewis, Nabila Ali, Amy Davies, Yvonne Wren, Kerry Humphries, Jonathan Sandy, Elliott Rees, Kimberley Marie Kendall, Gemma C Sharp, Michael J Owen, Marianne B M van den Bree, Evie Stergiakouli

## Abstract

Cleft lip and/or palate (CL/P) is the most common craniofacial congenital anomaly and has been associated with higher risk of neurodevelopmental and behavioural problems indicating potential shared genetic factors between CL/P and neurodevelopmental disorders. In this study, we aimed to determine the prevalence of neurodevelopmental copy number variants (CNV) in children with CL/P and their link to early developmental and behavioural problems. Using data from the Cleft Collective, the largest UK-based national cohort study of children with CL/P, we determined the rates of neurodevelopmental CNVs in children with CL/P comparing them to the general population, explored differences by cleft type and investigated risk of developmental delays and behavioural problems among those with CL/P and neurodevelopmental CNVs. Children with CL/P had a higher prevalence of neurodevelopmental CNVs than participants in four population-based samples (3.7% vs 2.3% in the Avon Longitudinal Study of Parents and Children (ALSPAC), 2.0% in Born in Bradford (BiB), 2.3% in Millenium Cohort Study (MCS), 1.7% in UK Biobank, ORs(95%CIs): ALSPAC= 1.56(1.18-2.06), BiB= 1.84(1.37-2.45), MCS= 1.59(1.19-2.11), UK Biobank= 2.15(1.68–2.71). Children with cleft palate only were 3 times more likely to have a neurodevelopmental CNV (95%CIs1.50-6.59, p=0.03) than children with cleft lip only. Furthermore, children with CL/P and neurodevelopmental CNVs were more likely to experience early developmental delays and behavioural problems by age 5 compared to children with CL/P and without neurodevelopmental CNVs. These findings highlight that genetic testing ascertaining the presence of neurodevelopmental CNVs might be helpful in early identification of developmental needs in children with CL/P.

## Introduction

Cleft lip and/or palate (CL/P) is the most common craniofacial congenital anomaly affecting one in every 1,000 live births (1). An estimated 30% of children born with CL/P have a syndromic form of cleft while for the remaining 70%, CL/P is considered non-syndromic. Syndromic CL/P usually presents with additional clinical phenotypes ranging from cognitive impairment to motor difficulties (2) and is caused by distinct genetic mutations or chromosomal differences (1). Similar to other multifactorial phenotypes, including neurodevelopmental and behavioural problems, non-syndromic CL/P has a complex aetiology with both genetic and environmental factors implicated in the causal pathways to phenotypes (1, 3).

The most common types of CL/P are known as cleft lip (CL), cleft lip and palate (CLP), cleft palate only (CP) and submucous cleft palate (where some of the soft, but not hard, palate fails to fuse properly). Cleft palate has long been considered aetiologically different to other types of cleft, with different genetic loci implicated from genetic (4) and epigenetic (5) studies as well as higher rates of syndromic forms (6).

Some studies have reported higher rates of neurodevelopmental and behavioural problems in children born with CL/P compared to children from the general population (7–10), educational underachievement (11, 12). A meta-analysis of behavioural problems in children born with CL/P reported that differences were mainly limited in higher rates of depression and anxiety symptoms(13). However, this meta-analysis was conducted before larger recent investigations. While environmental factors secondary to CL/P such as surgeries, appearance differences and bullying might be contributing to the increased prevalence of some behavioural problems among children born with CL/P (14), it is possible that the higher prevalence of neurodevelopmental and behavioural problems among children born with CL/P is partially explained by genetic overlap between CL/P and neurodevelopmental disorders.

Amongst the genetic variants implicated in CL/P and in neurodevelopmental and behavioural problems, copy number variants (CNVs) have emerged as significant contributors to pathogenesis. CNVs are a collective term for duplications and deletions in segments of chromosomal DNA that are greater than 1 kilobase (kb) in size. Specific CNVs have been shown to be associated with CL/P (15–17), neurocognitive impairment (18, 19) and complex neuropsychiatric disorders (20–25). Neurodevelopmental CNVs have not been systematically investigated in children born with CL/P, although there is some evidence of overlap between CNVs associated with CL/P and neurodevelopmental disorders, for example CP and developmental delay are both associated with 22q11 deletion syndrome (26). It is possible that rates of neurodevelopmental CNVs are higher in children born with CL/P compared to children from the general population. Given the heterogeneity in genetic factors involved in different types of cleft and particularly CP, it is likely that higher rates of neurodevelopmental CNVs are observed in certain types of cleft. Neurodevelopmental CNVs in children born with CL/P could be also linked to the higher rates of early developmental concerns(10, 27) and behavioural problems observed in children born with CL/P.(9)

We aim to determine the rate of neurodevelopmental CNVs among i) children born with any CL/P and ii) by specific cleft type, to describe which neurodevelopmental CNVs are present, and to test the following hypotheses:

(H1) Children born with CL/P have a higher rate of neurodevelopmental CNVs than the general population.

(H2) Rates of neurodevelopmental CNVs in children born with CL/P differ by cleft type.

(H3) Children born with CL/P and a neurodevelopmental CNV are at higher risk of developmental delay and behavioural problems than those born with CL/P and without a neurodevelopmental CNV. To assess early developmental problems in children born with CL/P we calculated mean developmental trajectories between 18 months to 5 years and compared them between children born with CL/P and a neurodevelopmental CNV and those born with CL/P but without a neurodevelopmental CNV. We also compared rates of behavioural problems between the two groups.

## Results

### Sample characteristics

The characteristics of the final sample of children born with CL/P from the Cleft Collective(28) who passed CNV Quality Control (QC) processes (N = 2180) are described in Table 1. The sample included more males (57%) than females (43%). Most participants had white mothers (91%), and 61% of all mothers had completed higher education (a degree or equivalent). Of those with genetic data for CNV calls, 998 (46%) had completed at least one neurodevelopmental or behavioural problem questionnaire during at least one-time point. A comparison of those with genetic data in the Cleft Collective to those without did not identify evidence of differences in terms of cleft type, syndromic status and mean scores in questionnaires assessing developmental delays and behavioral problems (S.Table 1).

**Table 1.**
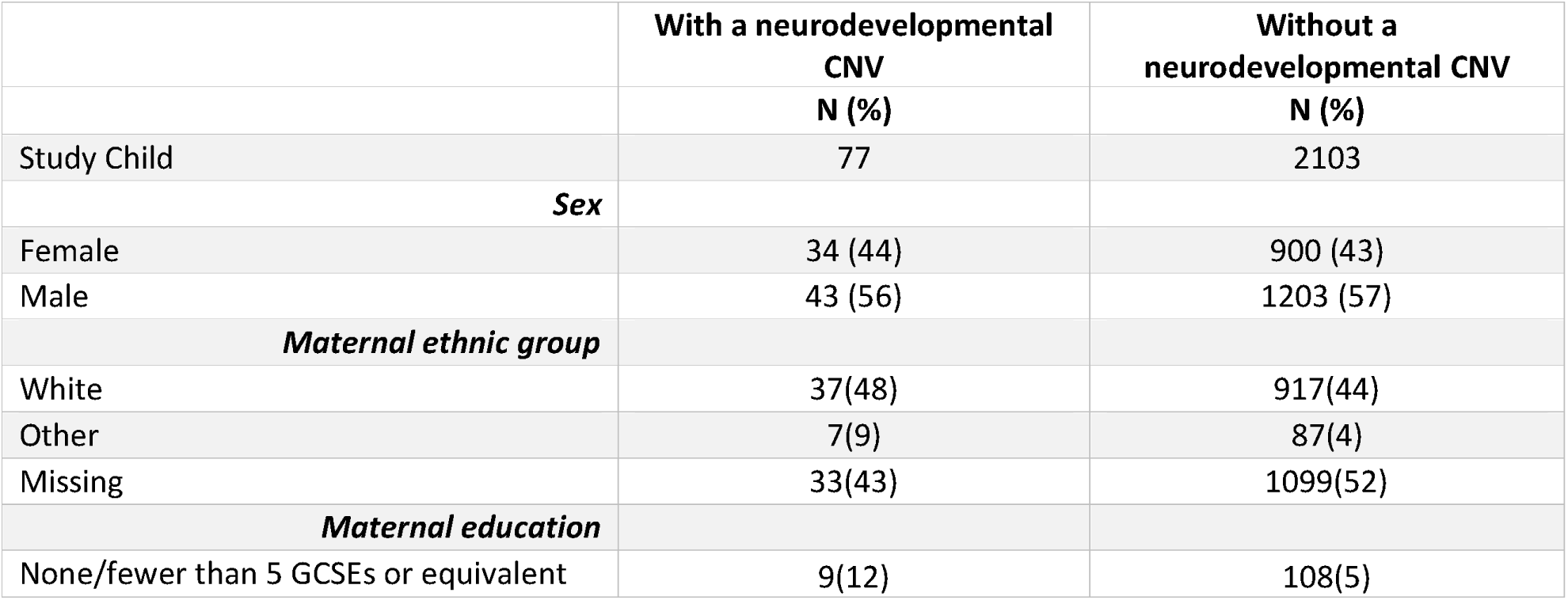
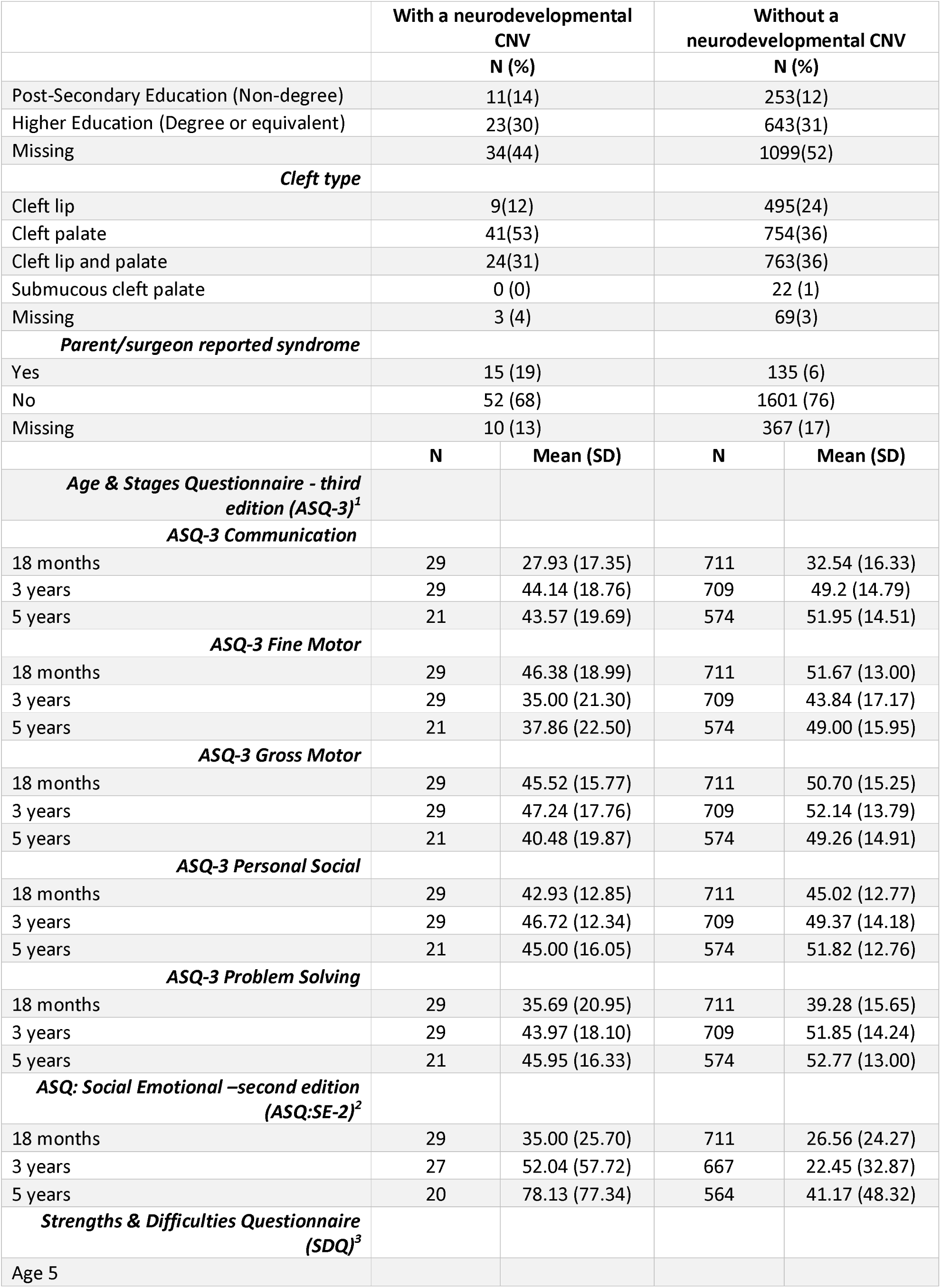
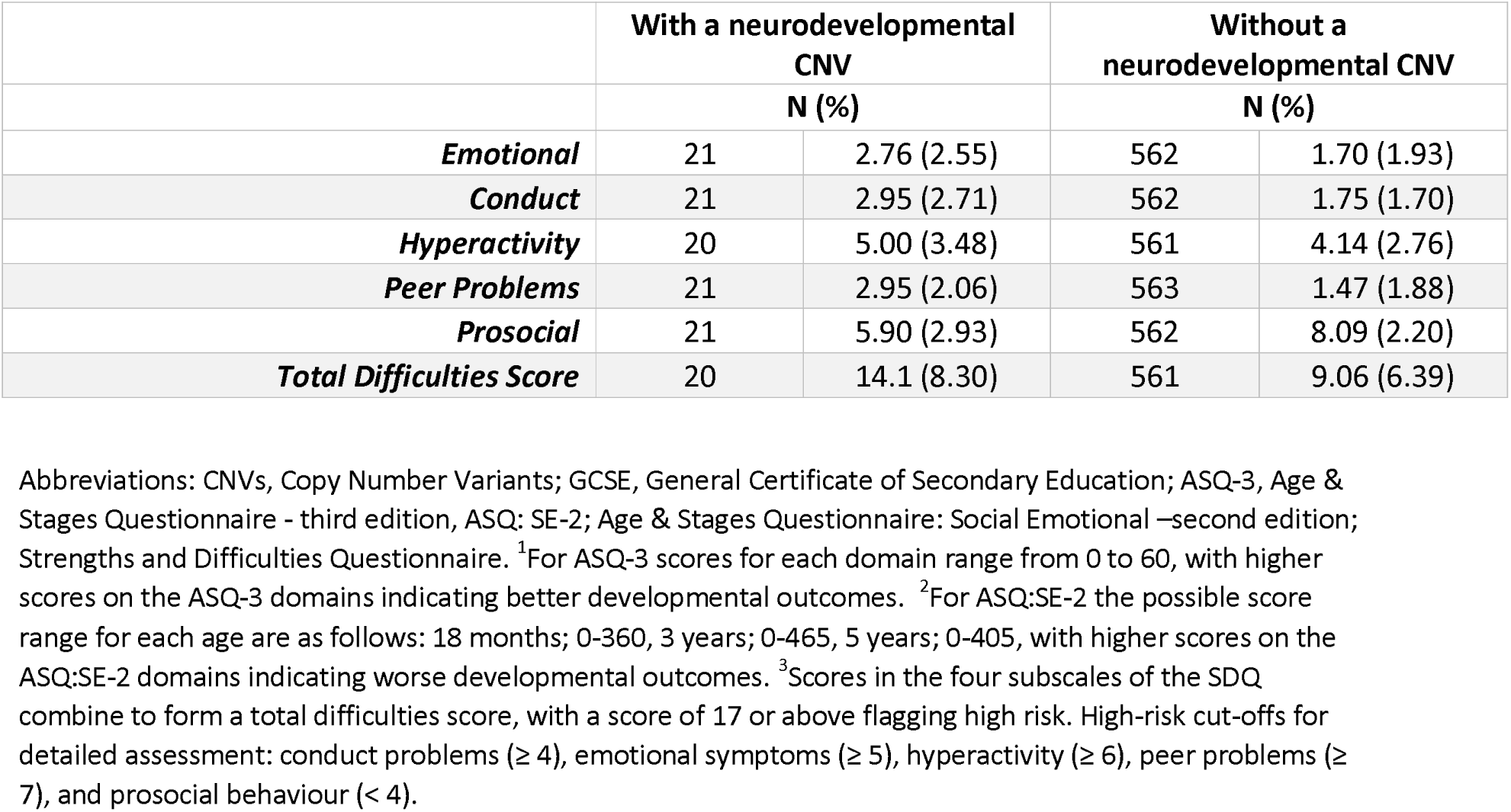
Sample Characteristics of Cleft Collective participants. Sample is defined as those with genetic and phenotypic data after quality control. Percentages calculated exclusive of missing data.

|  | With a neurodevelopmental CNV | Without a neurodevelopmental CNV |
| --- | --- | --- |
|  | N (%) | N (%) |
| Study Child | 77 | 2103 |
| Sex |  |  |
| Female | 34 (44) | 900 (43) |
| Male | 43 (56) | 1203 (57) |
| Maternal ethnic group |  |  |
| White | 37(48) | 917(44) |
| Other | 7(9) | 87(4) |
| Missing | 33(43) | 1099(52) |
| Maternal education |  |  |
| None/fewer than 5 GCSEs or equivalent | 9(12) | 108(5) |

|  | With a neurodevelopmental CNV |  | Without a neurodevelopmental CNV |  |
| --- | --- | --- | --- | --- |
|  | N (%) |  | N (%) |  |
| Post-Secondary Education (Non-degree) | 11(14) |  | 253(12) |  |
| Higher Education (Degree or equivalent) | 23(30) |  | 643(31) |  |
| Missing | 34(44) |  | 1099(52) |  |
| ***Cleft type*** |  |  |  |  |
| Cleft lip | 9(12) |  | 495(24) |  |
| Cleft palate | 41(53) |  | 754(36) |  |
| Cleft lip and palate | 24(31) |  | 763(36) |  |
| Submucous cleft palate | 0 (0) |  | 22 (1) |  |
| Missing | 3 (4) |  | 69(3) |  |
| ***Parent/surgeon reported syndrome*** |  |  |  |  |
| Yes | 15 (19) |  | 135 (6) |  |
| No | 52 (68) |  | 1601 (76) |  |
| Missing | 10 (13) |  | 367 (17) |  |
|  | N | Mean (SD) | N | Mean (SD) |
| ***Age & Stages Questionnaire - third edition (ASQ-3)<sup>1</sup>*** |  |  |  |  |
| ***ASQ-3 Communication*** |  |  |  |  |
| 18 months | 29 | 27.93 (17.35) | 711 | 32.54 (16.33) |
| 3 years | 29 | 44.14 (18.76) | 709 | 49.2 (14.79) |
| 5 years | 21 | 43.57 (19.69) | 574 | 51.95 (14.51) |
| ***ASQ-3 Fine Motor*** |  |  |  |  |
| 18 months | 29 | 46.38 (18.99) | 711 | 51.67 (13.00) |
| 3 years | 29 | 35.00 (21.30) | 709 | 43.84 (17.17) |
| 5 years | 21 | 37.86 (22.50) | 574 | 49.00 (15.95) |
| ***ASQ-3 Gross Motor*** |  |  |  |  |
| 18 months | 29 | 45.52 (15.77) | 711 | 50.70 (15.25) |
| 3 years | 29 | 47.24 (17.76) | 709 | 52.14 (13.79) |
| 5 years | 21 | 40.48 (19.87) | 574 | 49.26 (14.91) |
| ***ASQ-3 Personal Social*** |  |  |  |  |
| 18 months | 29 | 42.93 (12.85) | 711 | 45.02 (12.77) |
| 3 years | 29 | 46.72 (12.34) | 709 | 49.37 (14.18) |
| 5 years | 21 | 45.00 (16.05) | 574 | 51.82 (12.76) |
| ***ASQ-3 Problem Solving*** |  |  |  |  |
| 18 months | 29 | 35.69 (20.95) | 711 | 39.28 (15.65) |
| 3 years | 29 | 43.97 (18.10) | 709 | 51.85 (14.24) |
| 5 years | 21 | 45.95 (16.33) | 574 | 52.77 (13.00) |
| ***ASQ: Social Emotional –second edition (ASQ:SE-2)<sup>2</sup>*** |  |  |  |  |
| 18 months | 29 | 35.00 (25.70) | 711 | 26.56 (24.27) |
| 3 years | 27 | 52.04 (57.72) | 667 | 22.45 (32.87) |
| 5 years | 20 | 78.13 (77.34) | 564 | 41.17 (48.32) |
| ***Strengths & Difficulties Questionnaire (SDQ)<sup>3</sup>*** |  |  |  |  |
| Age 5 |  |  |  |  |
|  | N (%) |  | N (%) |  |
| ***Emotional*** | 21 | 2.76 (2.55) | 562 | 1.70 (1.93) |
| ***Conduct*** | 21 | 2.95 (2.71) | 562 | 1.75 (1.70) |
| ***Hyperactivity*** | 20 | 5.00 (3.48) | 561 | 4.14 (2.76) |
| ***Peer Problems*** | 21 | 2.95 (2.06) | 563 | 1.47 (1.88) |
| ***Prosocial*** | 21 | 5.90 (2.93) | 562 | 8.09 (2.20) |
| ***Total Difficulties Score*** | 20 | 14.1 (8.30) | 561 | 9.06 (6.39) |

Abbreviations: CNVs, Copy Number Variants; GCSE, General Certificate of Secondary Education; ASQ-3, Age & Stages Questionnaire - third edition, ASQ: SE-2; Age & Stages Questionnaire: Social Emotional –second edition; Strengths and Difficulties Questionnaire. <sup>1</sup>For ASQ-3 scores for each domain range from 0 to 60, with higher scores on the ASQ-3 domains indicating better developmental outcomes. <sup>2</sup>For ASQ:SE-2 the possible score range for each age are as follows: 18 months; 0-360, 3 years; 0-465, 5 years; 0-405, with higher scores on the ASQ:SE-2 domains indicating worse developmental outcomes. <sup>3</sup>Scores in the four subscales of the SDQ combine to form a total difficulties score, with a score of 17 or above flagging high risk. High-risk cut-offs for detailed assessment: conduct problems ( $\geq 4$ ), emotional symptoms ( $\geq 5$ ), hyperactivity ( $\geq 6$ ), peer problems ( $\geq 7$ ), and prosocial behaviour ( $< 4$ ).

#### Neurodevelopmental CNVs

We called a pre-determined list of 54 CNVs that have been previously associated with neurodevelopmental disorders (29) in children and parents from a UK-based national cohort study of children born with CL/P, the Cleft Collective, and compared their rates in four general population comparison groups; Avon Longitudinal Study of Parents and Children (ALSPAC) (30, 31), Born in Bradford (BiB) (32, 33), the Millenium Cohort Study (MCS) (34) and UK Biobank (35, 36) (see Methods for more details about the general population comparison groups used). The full list of CNVs used can be found in S.Table 2. After applying QC (described in Methods), we found strong evidence to support H1: Children born with any type of CL/P had a higher prevalence of neurodevelopmental CNVs than participants in all four general population comparison groups (3.7% vs 2.3% in ALSPAC, 2.0% in BiB, 2.3% in MCS, 1.7% in UK Biobank, ORs(95%CIs): ALSPAC = 1.56 (1.18-2.06), BiB = 1.84 (1.37-2.45), MCS = 1.59 (1.19-2.11), UK Biobank = 2.15 (1.68 – 2.71) (Table 2A and 2B). In addition, there was also strong evidence for H2: The prevalence of neurodevelopmental CNVs varied by cleft sub-type in children from the Cleft Collective (Table 2C). A total of 5.1% of children born with CP only had a neurodevelopmental CNV compared to 1.8% of children with cleft lip only. Children with CP were 2.98 times more likely to have a neurodevelopmental CNV (95%CIs= 1.50-6.59, p=0.03) compared to children with cleft lip only and children from the general population comparison groups (ORs = 2.3 – 3.18, in all general population comparison groups) (Table 2C). We also note that children with a neurodevelopmental CNV were more likely to have a syndromic cleft as reported by a parent or their surgeon (OR = 3.42, 95%CIs; 1.74, 6.36, p=2×10^-4^). However, 68% of those with neurodevelopmental CNVs did not report having a syndromic form of CL/P (Table 1).

**Table 2.** The frequency, percentage and odds ratios of children with neurodevelopmental disorder copy number variants (ND CNVs) among children with CL/P vs among general population cohorts and stratified by cleft type. Comparison cohorts are the Avon Longitudinal Study of Parents and Children (ALSPAC), Born in Bradford (BiB), the Millenium Cohort Study (MCS) and UK Biobank.

| Category | ND CNV+ | ND CNV- | Total | Percentage | OR | 95% CIs | p-value |
| --- | --- | --- | --- | --- | --- | --- | --- |
| CL/P | 77 | 2103 | 2180 | 3.7% | - | - | - |
| Cleft Lip (CL) | 9 | 495 | 504 | 1.8% | ref | ref | ref |
| Cleft Palate (CP) | 41 | 757 | 798 | 5.1% | 2.98 | 1.50-6.59 | 0.004 |
| Cleft Lip and Palate (CLP) | 24 | 767 | 791 | 3.0% | 1.72 | 0.79-3.73 | 0.17 |

B. CNV frequencies and rates in participants from the ALSPAC, BiB, MCS and UK Biobank.
| Category | ND CNV+ | ND CNV- | Total | Percentage |
| --- | --- | --- | --- | --- |
| ALSPAC | 144 | 6,217 | 6,361 | 2.3% |
| BiB | 149 | 7,477 | 7,626 | 2.0% |
| MCS | 151 | 6,559 | 6,710 | 2.3% |
| UK Biobank | 2,583 | 149,036 | 151,619 | 1.7% |

C. Comparisons of CNV rates between children born with CL/P, CL, CP, CLP from the Cleft Collective versus ALSPAC, BiB, MCS and UK Biobank.
| REFERENCE CATEGORIES |  |  |  |  |
| --- | --- | --- | --- | --- |
|  | ALSPAC | BiB | MCS | UK Biobank |
| Cleft Collective CL/P |  |  |  |  |
| OR | 1.56 | 1.84 | 1.59 | 2.15 |
| 95% CIs | 1.18 – 2.06 | 1.37- 2.45 | 1.19-2.11 | 1.68-2.71 |
| p-value | 0.002 | 3.9x10<sup>-5</sup> | 0.002 | 3.5x10<sup>-10</sup> |
| CL |  |  |  |  |
| OR | 0.77 | 0.91 | 0.79 | 1.07 |
| 95% CI | 0.34 – 1.52 | 0.41 – 1.79 | 0.35 – 1.55 | 0.48-2.05 |
| p-value | 0.535 | 1 | 0.637 | 0.729 |
| CP |  |  |  |  |
| OR | 2.3 | 2.72 | 2.35 | 3.18 |
| 95% CI | 1.57 – 3.31 | 1.86 – 3.9 | 1.61 – 3.37 | 2.26 – 4.37 |
| p-value | 1.2x10<sup>-5</sup> | 1.2x10<sup>-7</sup> | 5.8x10<sup>-6</sup> | 6x10<sup>-12</sup> |
| CLP |  |  |  |  |
| OR | 1.33 | 1.57 | 1.36 | 1.84 |
| 95% CI | 0.82 – 2.08 | 0.97 – 2.45 | 0.84 – 2.12 | 1.67 -2.76 |
| p-value | 0.21 | 0.056 | 0.17 | 1.6 x 10<sup>-6</sup> |

#### Neurodevelopmental CNVs and development trajectories

To assess early development in children born with CL/P, we used two questionnaires focusing on motor/communication development and emotional development. The Ages and Stages Questionnaire (ASQ-3) (37) assessing early life motor and communication development and the Ages and Stages Questionnaire - Social Emotional (ASQ:SE-2) (38) assessing early life social and emotional development were both completed by mothers when their children were at ages 18 months, 3 years, and 5 years. We calculated mean developmental trajectories of each domain in the ASQ-3 and the ASQ:SE-2 between 18 months to 5 years by fitting a series of linear multilevel models. Starting with a null model for each subscale we incrementally included random intercepts and slopes, sex, the interaction between age and sex, CNV status and cleft sub-type as covariates testing how each term impacted the fit of the data to the model. The terms that were included for each subscale are listed in S.Table 3. This analysis supports the hypothesis that people born with CL/P and a neurodevelopmental CNV are at higher risk of developmental delays, see Figure 1, than their peers born with CL/P alone (H3). In linear multilevel models of the ASQ-3 subscales, developmental trajectories of children with neurodevelopmental CNVs were estimated to be consistent with the developmental deficit hypothesis (39). In line with this hypothesis, developmental delays manifested early in life (lower intercept relative to peers without neurodevelopmental CNVs) but the rate of development (slope of trajectories) was the same as in peers without neurodevelopmental CNVs see Figure 1 and S.Table 3. However, children with neurodevelopmental CNVs did not catch up with their peers without CNVs by the age of 5 years.

**Figure 1:**
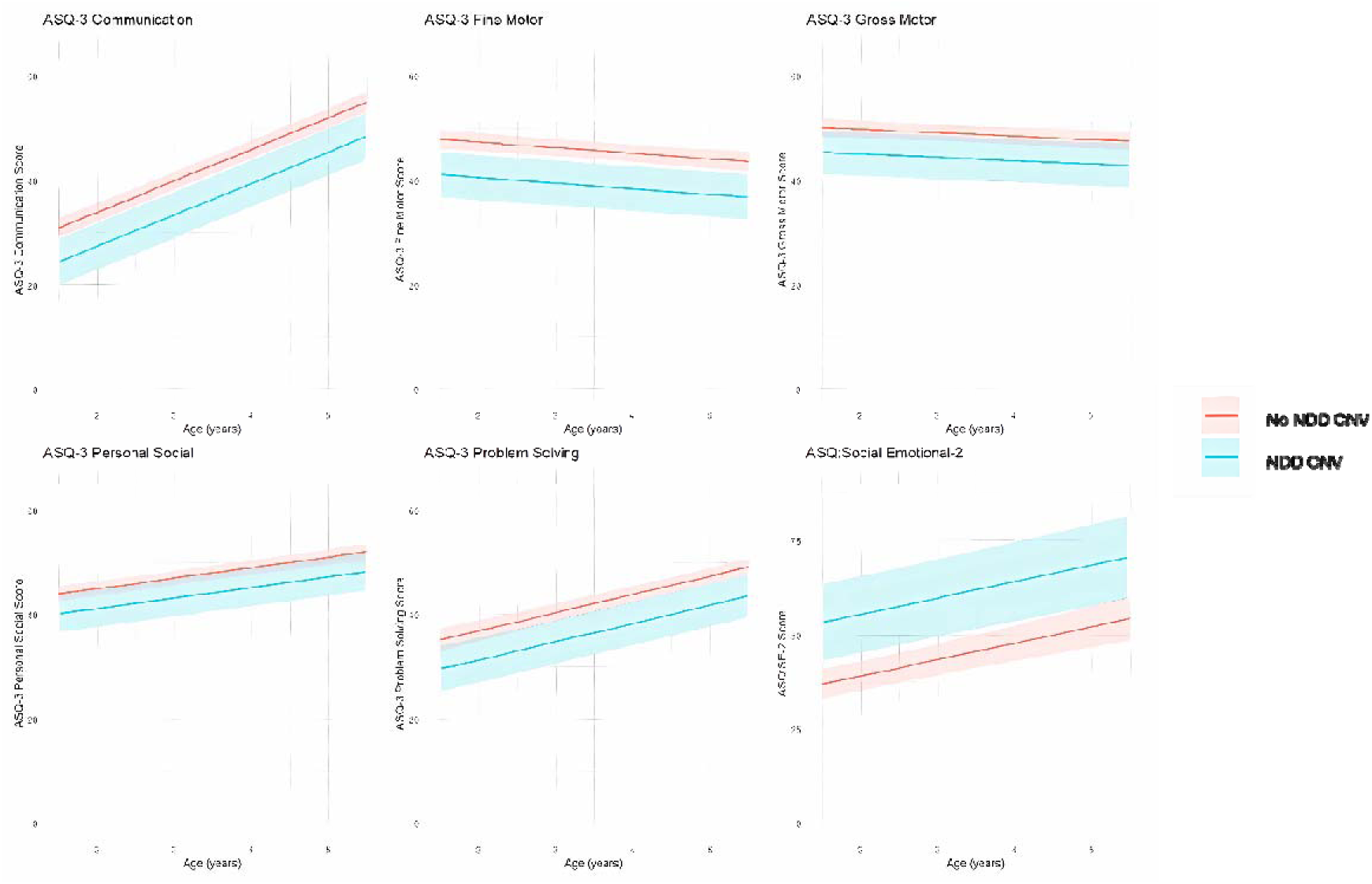
The best-fitting model of predicted scores and 95% confidence intervals from linear multilevel models of Ages and Stages Questionnaire (ASQ)-3 and ASQ:SE-2 scores from ages 18 months to 5 years. Plotted average scores are for a male with a cleft palate both with (blue) and without (red) neurodevelopmental copy number variants (neurodevelopmental CNVs). Please note higher scores on the ASQ-3 domains indicate better developmental outcomes and higher scores on ASQ:SE-2 indicate poorer social emotional outcomes.

**Table 3:** Fixed effect estimates of having a neurodevelopmental copy number variant (ND CNV) in linear multilevel models of the Ages and Stages Questionnaire (ASQ-3) and the Ages and Stages Questionnaire - Social Emotional (ASQ:SE-2) in children born with a CL/P. The reference category is no ND CNV. Higher scores on the ASQ-3 domains indicate better developmental outcomes and higher scores on ASQ:SE-2 indicate poorer social-emotional outcomes. Where parameters improved model fit, models are adjusted for sex, interaction with age and sex and cleft sub-type.

| *Development 18m to 5 years* | **ND CNV**<br>**N** | **No ND**<br>**CNV**<br>**N** | **$\beta$ - Estimate** | **95% Confidence**<br>**Intervals** | **P-**<br>**value** |
| --- | --- | --- | --- | --- | --- |
| **ASQ-3 Communication** | 38 | 941 | -6.52 | -10.83, -2.22 | 0.003 |
| **ASQ-3 Fine Motor** | 38 | 938 | -6.80 | -10.98, -2.62 | 0.001 |
| **ASQ-3 Gross Motor** | 38 | 940 | -4.71 | -8.76, -0.65 | 0.023 |
| **ASQ-3 Personal Social** | 38 | 939 | -3.80 | -7.25, -0.35 | 0.031 |
| **ASQ-3 Problem Solving** | 38 | 939 | -5.56 | -9.58, -1.54 | 0.007 |
| **ASQ: Social Emotional-2** | 37 | 930 | 16.41 | 6.52, 26.29 | 0.001 |

Each ASQ:SE-2 score from 18 months through to 5 years indicated more developmental impairment across all domains among those with any CL/P and neurodevelopmental CNVs. Those with a neurodevelopmental CNV were estimated to score 16.41 points higher (higher scores on the ASQ:SE-2 indicate higher risk of social-emotional impairment) on the ASQ:SE-2 total score (95%CI 7.05, 26.98, p = 4×10^-11^) at 18 months and this persisted through development to age 5 indicating a stable trend of worse social-emotional development relative to peers with CL/P who do not have an neurodevelopmental CNV (Table 3).

Cleft palate only, compared to cleft lip only, was associated with poorer outcomes across all ASQ-3 developmental domains at each time point (see S.Table 4). Children with submucous cleft palate also showed larger negative effects across ASQ-3 domains compared to children with cleft lip only, though confidence intervals were wide due to the small sample size of children with submucous cleft palate. Similarly, ASQ:SE-2 scores indicated more social-emotional problems for children with CP only and submucous cleft palate compared to those with cleft lip only, though wide confidence intervals were also noted for the submucous cleft palate group.

#### Neurodevelopmental CNVs and behavioural outcomes

We tested if children born with CL/P and neurodevelopmental CNVs were at higher risk of behavioral problems at age 5 as indicated by the Strengths and Difficulties Questionnaire (SDQ) (40) compared to children with CL/P alone by using linear regression models. Children with CL/P and a neurodevelopmental CNV were estimated to score 4.21 points higher (95%CI 1.48, 6.94, p= 0.003) on the SDQ total difficulties scale than their peers with CL/P and no neurodevelopmental CNV. At age 5, children with CL/P and neurodevelopmental CNVs scored higher on average across conduct, emotional, hyperactivity and peer problem subscales and lower on average on the prosocial subscale, all indicating a larger proportion of individuals with higher rates of behavioral difficulties in this group relative to peers with CL/P and no neurodevelopmental CNV, see Table 4. Pooled estimates from imputed data were similar to estimates from the complete cases analyses reported in S.Table 5.

**Table 4.** Estimated pooled effect of having a neurodevelopmental disorder copy number variant (ND CNV) in linear regression models of the Strengths and Difficulties Questionnaire (SDQ) at age 5. The reference category is no ND CNV. Higher scores on the SDQ indicate higher risk of behavioural problems except for the prosocial subscale where higher scores indicate more prosocial traits. Multiple imputation was used to manage missing data in the SDQ.

| *SDQ domains at 5 years* | ND CNV<br>N | No ND<br>CNV<br>N | **Pooled**<br>**effect** | **95% Confidence**<br>**Interval** | **P-value** |
| --- | --- | --- | --- | --- | --- |
| **Conduct** | 39 | 959 | 1.00 | (0.24, 1.77) | 0.01 |
| **Emotional** | 39 | 959 | 0.92 | (0.13, 1.72) | 0.023 |
| **Hyperactivity** | 39 | 959 | 0.52 | (-0.63, 1.68) | 0.414 |
| **Peer Problems** | 39 | 959 | 1.13 | (0.38, 1.87) | 0.003 |
| **Prosocial** | 39 | 959 | -1.89 | (-2.85, -0.93) | 0.0001 |
| **SDQ Total** | 39 | 959 | 4.21 | (1.48, 6.94) | 0.0025 |

SDQ analyses by cleft subtype did not indicate any evidence of differences in behavioural problems by cleft subtype, although sample sizes for this analysis were very limited (see S.Table 6).

#### Individual Neurodevelopmental CNV Loci

Table 5 summarizes the frequencies of specific neurodevelopmental CNV loci identified > 10 times in children with CL/P compared to the four general population comparison groups we used, with details on all the CNV loci identified in S.Table 7. To protect anonymity and comply with ethics regulations, individual loci in each cohort where N<5 are not shown. Three loci—15q11.2, 22q11.2, and 16p11.2—accounted for 73% of the 77 neurodevelopmental CNVs identified. The rates of 15q11.2 deletions and duplications were similar to estimates from the general population, while the 22q11.2 deletion rates were, as expected, elevated in the Cleft Collective (22q11 deletion syndrome is associated with higher risk of cleft palate(26)). We also found a higher proportion of 16p11.2 deletions (0.46%) among children with CL/P, despite this CNV being extremely rare in the general population (<0.03% in UK Biobank and <5 cases in the other cohorts).

**Table 5:** Neurodevelopmental copy number variation in specific loci among children with cleft vs general population samples. The 4 loci included are those where. 10 or more children were identified with that neurodevelopmental CNV among children in the Cleft Collective. To protect anonymity and comply with ethics regulations, individual loci in each cohort where N<5 are not shown.

|  |  | 15q11.2 del BP1-BP2 |  |  |  | 15q11.2 dup BP1-BP2 |  |  |  | 16p11.2 del |  |  |  | 22q11.2 del |  |  |  |
| --- | --- | --- | --- | --- | --- | --- | --- | --- | --- | --- | --- | --- | --- | --- | --- | --- | --- |
|  |  | N (%) | OR | 95% CI | P-value | N (%) | OR | 95% CI | P-value | N (%) | OR | 95% CI | P-value | N (%) | OR | 95% CI | P-value |
|  | **Cleft Collective**<br>**(N = 2,180)** | 17<br>(0.78) |  |  |  | 13<br>(0.60) |  |  |  | 10<br>(0.46) |  |  |  | 10<br>(0.46) |  |  |  |
| **Reference**<br>**Categories** | **ALSPAC**<br>**(N = 6,217)** | 42<br>(0.68) | 1.16 | 0.62,<br>2.08 | 0.655 | 32<br>(0.51) | 1.16 | 0.56,<br>2.28 | 0.613 | <5 | - | - | - | <5 | - | - | - |
|  | **BiB**<br>**(N = 7,477)** | 29<br>(0.39) | 2.02 | 1.04,<br>3.81 | 0.032 | 44<br>(0.59) | 1.01 | 0.50,<br>1.92 | 1 | <5 | - | - | - | <5 | - | - | - |
|  | **MCS**<br>**(N = 6,559)** | 52<br>(0.79) | 0.98 | 0.53,<br>1.73 | 1 | 38<br>(0.58) | 0.92 | 0.45,<br>1.77 | 0.875 | <5 | - | - | - | <5 | - | - | - |
| | **UK Biobank**<br>**(N = 151,619)** | 544<br>(0.36) | 2.18 | 1.26,<br>3.54 | 0.004 | 762<br>(0.50) | 1.19 | 0.63,<br>2.05 | 0.539 | 43<br>(0.03) | 16.24 | 7.27,<br>32.91 | $4.7 \times 10^{-13}$ | 5<br>(0.00) | 139.29 | 43.46,<br>529.08 | $9.8 \times 10^{-15}$ |

#### Neurodevelopmental CNVs in relatives of children with cleft

We derived neurodevelopmental CNVs in parents and siblings of children from the Cleft Collective. Neurodevelopmental CNVs were present in 2.3% of mothers (54/2,344), 3.0% of fathers (54/1,869), and 3.4% of siblings (5/144) of children born with CL/P. These rates were similar to those found in general population comparison groups (ALSPAC, BiB, MCS) (S. Table 8). There was some evidence that parents of children with CL/P had higher odds of having a neurodevelopmental CNV compared to adults in the UK Biobank but not compared to other general population comparison groups (see S.Table 8).

#### Inherited vs *de novo* Neurodevelopmental CNVs

We determined if CNVs in children from the Cleft Collective were inherited or *de novo* where parental CNV data were available. Of the 77 children with neurodevelopmental CNVs, 31 (40%) inherited them from a parent, 19 (25%) were *de novo*, and 27 (35%) had an unknown origin. Out of the 27 Individuals with an NDD CNV of unknown origin, 12 had neither parent present in the genotypic data, 8 had only their mother genotyped, 6 had only their father genotyped, and one had both parents whose genotypic data failed QC checks. We investigated whether there were any differences in ASQ-3 and ASQ:SE-2 scores by whether the CNVs arose *de novo* in the child or whether they were inherited from the parents. Whilst there was little to no evidence that *de novo or inherited* neurodevelopmental CNVs impacted early development, there were strong negative associations with developmental outcomes for children who did not have both parents provide genetic data, hence had unknown origin of their neurodevelopmental CNV, see S.Table 9. Due to small sample sizes, we were not able to test for differences in SDQ scores by inherited or *de novo* CNV status.

## Discussion

This is the first study to investigate and to identify that neurodevelopmental CNVs are more common among children born with CL/P (3.7%) than in the general population (∼ 2.2%) (H1). Further, we identified that neurodevelopmental CNVs are most common among the subgroup of children born with a CP only (H2). Making use of longitudinal data we observed that neurodevelopmental CNVs are associated with early development deficits and behavioural problems among children born with CL/P from 18 months to 5 years of age (H3). In addition, we observed that along with 22q11, 16p11.2 appears to be a specific CNV locus relevant to CP.

Children with CL/P are more likely to have neurodevelopmental CNVs compared to the general population, and the evidence indicates that those with both CL/P and neurodevelopmental CNVs face a higher risk of behavioural problems and developmental delays than their peers with CL/P but without neurodevelopmental CNVs. These findings suggest that neurodevelopmental CNVs may have pleiotropic effects, influencing both the development of CL/P and behavioural differences. Alternatively, children with CL/P and neurodevelopmental CNVs may have more complex medical needs and/or be more likely to be exposed to adverse experiences, such bullying and discrimination, compared to their peers with CL/P but no neurodevelopmental CNVs which in turn could be increasing behavioural symptoms. Regardless of the specific causal pathway, CNVs that increase risk to neurodevelopmental disorders are observed in higher rates in children born with CL/P compared to children from the general population and this could be linked to their higher rates of behavioural problems and developmental delays. However, the majority of children with a neurodevelopmental CNV in the Cleft Collective (68%) were not recorded as having a syndromic form of CL/P (from parent or surgeon reports). This suggests that the majority of parents are probably unaware of their child’s CNV status and the health implications this might have currently or in the future.

Children born with CP only were more likely to have neurodevelopmental CNVs and developmental delays compared to children born with cleft lip only. This is consistent with previous research on CP indicating higher risk of having a syndromic cleft amongst those with CP only. In addition, genome-wide association studies and epigenome-wide association studies have identified CP as a phenotype with distinct aetiology from cleft lip and other cleft phenotypes (4, 5, 41).

Rates of deletions in the 16p11.2 locus were unusually high among the Cleft Collective children; 10 in 2,180 (0.46%) as compared to general population estimates of 3 in 10,000 (43/151,619, 0.03%) in UK Biobank and less than five cases in ALSPAC, BiB and MCS. Recent work on 16p11.2 deletion syndrome suggests that a minor proportion of those affected present with small craniofacial malformations, among which cleft is mentioned, but the core distinguishing features for the majority with this neurodevelopmental CNV are developmental delays in language, motor difficulties, autism, seizures and obesity in adulthood (19, 25, 42, 43). Communication and language are also commonly delayed across 15q.11.2 (44) and 22q.11.2 (26) deletion syndromes.

The interpretation of impaired communication among children born with CL/P is not straightforward. Low scores in the communication subscale may represent a causal effect of CL/P (45) and not a direct genetic effect, i.e. children born with CLP or CP have a higher risk of speech disorder which in turn can impact language and communication development. Nevertheless, those with neurodevelopmental CNVs and CL/P did consistently score lower than their peers in the Cleft Collective on communication subscales, perhaps indicative of a ‘double hit’ on the development of communication among those with CL/P and neurodevelopmental CNVs.

Recent longitudinal research has described the development of children with non-syndromic CL/P as delayed on each of the ASQ-3 subscales relative to typically developing children and differences emerged between 18 months and 2 years (10). In the present study, those with CL/P and neurodevelopmental CNVs showed early deficits on the ASQ-3 compared to those with CL/P but without neurodevelopmental CNVs, although the rate of development was the same in both groups. These differences were stable and persisted from 18 months through to 5 years across each subscale in line with the developmental deficit hypothesis (39). According to this hypothesis, impaired development manifests early in life. Despite the rate of development being the same as in typically developing children, children with CL/P and neurodevelopmental CNVs do not catch up from this early developmental deficit. Thus, while those without neurodevelopmental CNVs may also be at risk of delayed development, our data suggest that the vulnerability of those with CL/P and neurodevelopmental CNVs is greater. Findings from a study of children with 22q11.2 deletion syndrome, which has cleft palate as one of its associated features, and unaffected siblings also supported the developmental deficits hypothesis. There was little evidence of deterioration in trajectories of cognitive development in children with 22q11.2 deletions despite the early deficits compared to unaffected siblings (46).

Given that a proportion of neurodevelopmental CNVs are inherited, it is possible that indirect genetic effects may play a role i.e. that parental genetic architecture impacts parenting resources and the child’s early environment hence later development. However, in this study, we did not find evidence to suggest that inherited or *de novo* CNVs had a greater impact on outcomes. Rather, we found that the ’unknown’ category had strong evidence of association with early development and behavioural outcomes. This suggests that having two parents participate and providing genetic data seems protective for developmental delay. In other words, selection bias into the study had a stronger effect than whether a neurodevelopmental CNV was inherited or *de novo*.

### Strengths and Limitations

The main challenge of examining cleft, a rare outcome, in the context of rare genetic features (CNVs), is having a sufficient sample size to detect effects. The Cleft Collective is the largest longitudinal dataset of children with CL/P with genotypes available on children and their parents. The effects reported in this study are sufficiently large that even where samples sizes are limited, we were able to observe strong associations. This serves to support the overall conclusion that even with small numbers we can observe that neurodevelopmental CNVs are more common among children with cleft and have a notable impact on both development and behavioural problems.

Making use of four different general population samples each with their own different selection biases (47) and consistently estimating similar results across these samples adds strength to the confidence we can have in our results. Specifically, ALSPAC, BiB and MCS are birth cohorts and comparable in terms of data collection methodology and timings to the Cleft Collective, which for genetic data was early in life. UK Biobank provides a much larger sample, but recruited 40 – 69-year-olds and hence estimates of neurodevelopmental CNVs are anticipated, and are observed, to be lower due to survival bias. However, this large sample allows for comparisons even for rare outcomes, such as 16p.11.2 deletions.

Selection bias and attrition are important concerns in any longitudinal study and they are associated with genetic liability to neurodevelopmental and psychiatric phenotypes(48, 49). We observed selection bias in the Cleft Collective with strong associations of parents not providing DNA with lower developmental outcomes for children. This suggests that we might be underestimating the rates of neurodevelopmental CNVs in the Cleft Collective if parents of children with neurodevelopmental concerns and/or CNVs are less likely to participate.

We note that we only examined one type of rare genetic mutations. Common variants have long been established to be associated with CL/P (4, 50) and neurodevelopmental disorders (51–53) and examining whether they contribute to behavioral problems in children born with CL/P is equally important.

There are several statistical tests performed in this analysis which does increase the chance of type 1 error, however, we present confidence intervals around all our effect estimates and urge the reader to infer the robustness of an effect estimate using these values as opposed to applying a multiple testing correction and shifting arbitrary p-value thresholds(54, 55).

Clinical diagnoses of specific neurodevelopmental disorders were not assessed in this study. Instead, we relied on scores measured by scales, such as SDQ and ASQ which are strongly predictive of developmental and behavioural problems (38, 40, 56). The study population would be too young by age 5, to have received a diagnosis of a childhood neurodevelopmental or psychiatric disorder, hence it is not appropriate to use categorical measures. Further, the use of continuous scores maximises variance in the data to provide greater statistical power. The early indicators of developmental and behavioural problems that we identified do however provide a strong rationale for continued follow up in the Cleft Collective and/or linkage with primary and secondary care studies.

### Implications

The evidence set out in this paper suggests children born with CL/P are at higher risk of neurodevelopmental CNVs (3.7% vs 2.2% in the general population), and children with neurodevelopmental CNVs and CL/P are at higher risk of behavioural problems at 5 years and developmental delays before the age of 5 years. The implication is that genetic testing to ascertain the presence of neurodevelopmental CNVs might be helpful in early identification of developmental needs in children born with CL/P, as well as signposting the need for follow up and early interventions (57) to reduce the impact as children grow. This is particularly important because the majority of parents whose child had a CNV did not report being aware that they might have a syndromic form of cleft. One of the strongest associations noted in this study was the social-emotional development assessed by ASQ:SE-2 scores which include parent-reported concerns about their child. Contextualizing parental concern with possible explanatory power of genetic testing may help support parents understanding their child’s developmental needs. The results in this paper also substantiate the policy among cleft teams of the inclusion of clinical psychologists to support children born with CL/P.

## Conclusion

Children born with CL/P are at greater risk of carrying an neurodevelopmental CNV than the general population, with the highest risk observed in those with CP only. Furthermore, those with both an neurodevelopmental CNV and CL/P experience an elevated risk of early developmental delays from 18 months to 5 years and are more likely to experience behavioural problems by age 5. These findings underscore the importance of early genetic screening and tailored interventions to address the unique developmental challenges faced by this group. Follow-up of children’s developmental and behavioural traits as they grow up as well as linkage to their educational attainment data could reveal further consequences of neurodevelopmental CNVs in children born with CL/P.

## Materials and Methods

### Cleft Collective

The Cleft Collective(28) is an ongoing UK-based national cohort study of children born with CL/P, their parents, and siblings. Details of recruitment and data collection procedures can be found elsewhere (8, 9) along with how to access the resource at https://www.bristol.ac.uk/cleft-collective/professionals/access/. Briefly, data are collected from two cohorts, the birth cohort and a cohort of five-year olds. Families are recruited to the birth cohort during pregnancy (if cleft is detected during ultrasound scans) or soon after birth but before the study child’s primary surgery to repair their cleft. Families are recruited to the five-year cohort between the child’s fifth and sixth birthday, often at a five-year follow-up clinic. Data used to address the outlined hypotheses are from a combination of parental questionnaires and biological samples collected by families or medical professionals. All study participants were recruited with informed parental consent. The Cleft Collective Cohort Studies received ethical approval from the South-West Central Bristol NRES Ethics Committee (REC 13/SW/0064). This project was approved by the Cleft Collective project management group (CC048-ES).

### Genotyping

Study children from the birth cohort provided blood and/or tissue samples, consenting parents and siblings from both cohorts and study children from the 5-year-old cohort provided saliva samples. These were all genotyped in the Illumina facility in Bristol Bioresource Laboratories, Bristol UK, using the Illumina Global Screening Array (GSA) version 3. Raw data from 7182 samples were uploaded to GenomeStudio where we carried out preliminary Quality Control (QC) processes in which 264 samples were removed, see S.Figure 1. We exported log R ratios and B-allele frequencies for each of the remaining 6918 samples to identify CNVs.

### CNV calling

We limited the CNVs called to a pre-determined list of 54 CNVs that have been associated with neurodevelopmental disorders (29), the full list is reported in S.Table 2. We adapted the following neurodevelopmental CNV calling pipeline: https://github.com/CardiffMRCPathfinder/NeurodevelopmentalCNVCalling.git (58) using PennCNV 1.0.5 (59). This is described in detail in the Supplementary Note 1 and S.Figure 2. Briefly, we performed initial QC excluding samples based on 100 or more CNVs, waviness factor < -0.037 or >0.037, or log R ratio SD > 0.24. From 7182 raw samples, the final sample consisted of 6551, see S.Figure1, S.Figure 3 and S.Figure4. We visually inspected log R ratios and B-allele frequencies of each NDD CNV to exclude low quality calls, see S.Figure 5 for examples of CNV calls.

Where data were available for a study child and both parents, we were able to determine if a neurodevelopmental CNV was inherited or *de novo*. Where genetic data from either parent were missing, inherited/*de novo* status remained unknown.

### Neurodevelopmental CNV control samples

We included published rates of neurodevelopmental CNVs in children from the Avon Longitudinal Study of Parents and Children (ALSPAC)(60), Born in Bradford (BiB)(33) , the Millenium Cohort Study (MCS) (60) and adult data from UK Biobank (36) to provide a general population baseline for neurodevelopmental CNVs. ALSPAC is a long-standing multi-generational cohort, recruited in the early 1990s from prospective mothers in the county of Avon and has multiple waves of rich phenotypic information on them and their offspring(30, 31). BiB was established in Bradford in 2007, recruiting prospective mothers, and has notable ethnic diversity compared with other UK-based cohorts(32). MCS is a nation-wide cohort from children born in 2000-2001, with intentional oversampling of areas with ethnic minorities as well as deprivation(34). UK Biobank is a large-scale database of over 500,000 UK adults providing extensive genetic, lifestyle, and health information (35).

For each cohort the same set of 54 neurodevelopmental CNVs were identified and the same pipeline used to perform the calls (29).

### Measures

#### Demographics

We estimated sex from genetic data using the ‘sex estimate’ function in GenomeStudio2.0. Maternal ethnicity and educational attainment were collected via baseline questionnaires. To ensure participant anonymity and comply with our ethical approval which stipulates cell counts ≥ 5, six ethnic groups were collapsed to two, ‘white’ and ‘other’. Maternal education categories were collapsed from 14 options into three groups, ‘None/fewer than 5 GCSEs or equivalent’, ‘Post-Secondary Education (Non-degree)’ and ‘Higher Education (Degree or equivalent)’.

#### Cleft sub-type

Cleft sub-type (cleft lip, cleft palate, unilateral cleft lip and palate, bilateral cleft lip and palate, and submucous palate) was derived from parent-reported and surgeon-reported questionnaires. Due to low cell counts, we combined the ‘unilateral’ and ‘bilateral’ categories to form a new category ‘cleft lip and palate’. Parent/surgeon-reported syndrome was derived from a binary variable from surgeon and/or parental questionnaires.

#### Neurodevelopmental and behavioural problem measures

All measures below were administered in the form of questionnaires completed by both mothers and fathers.

## Developmental delays

The Ages and Stages Questionnaire (ASQ-3) (37) was used to assess development across five domains: Communication, Gross Motor, Fine Motor, Problem Solving, and Personal-Social. Each domain is evaluated through six items, where responses are categorised as ‘yes’ (10 points), ‘sometimes’ (5 points), or ‘not yet’ (0 points). Scores for each domain range from 0 to 60, with higher scores indicating more advanced development. In cases where up to two items were missing within a domain, these missing answers were replaced with the mean of the items that were answered. The ASQ-3 was administered at ages 18 months, 3 years, and 5 years. For reference, the cut offs for each ASQ measure which indicate clinical monitoring are outlined in S.Table 10.

## Social and Emotional Development

Social and emotional development was measured using the Ages and Stages Questionnaire - Social Emotional (ASQ:SE-2) (38). Higher scores reflect poorer social-emotional functioning. The scores are treated as continuous variables for analysis. The ASQ:SE-2 was collected at ages 18 months, 3 years, and 5 years. For reference, the cut-offs for each ASQ measure which indicate clinical monitoring or referral are outlined in S.Table 10. The possible score range for each age is as follows: 18 months; 0-360, 3 years; 0-465, 5 years; 0-405.

## Behavioural problems

The Strengths and Difficulties Questionnaire (SDQ) (40), administered at age 5, was used to assess behavioural problems. This 25-item screening tool covers five subscales: conduct problems, emotional symptoms, hyperactivity, peer problems, and prosocial behaviour. There are 5 items on each subscale and options for each item are ‘not true’ coded as zero, ‘somewhat true’ coded as one and ‘certainly true’ coded as two. Higher scores on all subscales, except prosocial behaviour, indicate a higher risk of behavioural issues. Four subscales (conduct problems, emotional symptoms, hyperactivity, peer problems) combine to form a total difficulties score, with a score of 17 or above flagging high risk. High-risk cut-offs for detailed assessment are: conduct problems (four or more), emotional symptoms (five or more), hyperactivity (six or more), peer problems (seven or more), and prosocial behaviour (less than four) (61).

For each of the neurodevelopmental and behavioural measures described above we used continuous scores to maximise variance in the data (56). Due to a higher response rate, maternal responses were the default score used; where maternal data were unavailable, we used paternal data to complete scores. Correlations between the equivalent scores from maternal and paternal raters were positive and high across all SDQ measures, low to moderate for ASQ:SE-2 and moderate to high for all ASQ-3 measures, see S.Figures 6 and 7.

### Statistical analysis

To test H1, we generated frequency tables with counts of Cleft Collective study children with and without neurodevelopmental CNVs, and the equivalent reference data from four general population comparison groups: Avon Longitudinal Study of Children and Parents (ALSPAC), Millenium Cohort Study (MCS), Born in Bradford (BiB) and UK Biobank. We calculated odds ratios (ORs), confidence intervals, and p-values using Fischer’s exact test. We repeated these analyses stratified by cleft type (H2). To further assess the prevalence of neurodevelopmental CNVs by cleft type we performed logistic regression with presence of neurodevelopmental CNVs as an outcome, cleft sub-type as a categorical predictor and sex as a covariate.

### Multilevel models

To determine the best fit of mean development of each domain in the ASQ-3 and the ASQ:SE-2 across the three time points, between 18 months to 5 years, we fitted a series of linear multilevel models increasing in complexity from null models to random intercepts and random slopes. We incrementally included sex, the interaction between age and sex, and cleft sub-type as covariates. The interaction term was included to model how sex-specific trajectories change over time. Covariates were included in the model if they improved model fit by reducing Akaike Information Criterion (AIC) and showing a Loglikelihood Ratio (LR) tests at P < 0.05. Age was centered at 18 months. The fit statistics and final formula for each model are presented in S.Table 3.

To test H3, we included the presence or absence of a neurodevelopmental CNV as a level-1 predictor in each multilevel model. We also tested for any interactions between neurodevelopmental CNVs and age to assess if there were changes in the impact of neurodevelopmental CNVs over time. To assess if subgroups of children with neurodevelopmental CNVs develop differently we included ‘cleft sub-type’ and ‘inherited vs de novo’ as additional analyses. All multilevel models were estimated using ‘R2MLwiN’ in R and ‘MLwiN’(62, 63).

To further test H3 in a cross-sectional framework and estimate associations between the presence of a neurodevelopmental CNV and SDQ scores at age 5 we used linear regression models. In each regression model we included sex and age as covariates, we retained the covariates where LR test at P <0.05 indicated improved fit. All statistical analyses were performed in R 4.3.2.

### Missing data

We used multiple imputations to manage missing data in linear regression models. The imputation model was limited to those who had at least one outcome measure (ASQ-3, ASQ:SE-2 or SDQ) present at one time point. We included all outcome measures at each time point (ASQ-3, ASQ:SE-2, SDQ) and all demographic measures listed above. We used the package “mice”(64) in R and having established the percentage of missing data was ∼30%, generated 30 complete datasets through 10 iterations of predictive mean matching. We report on the overall pooled estimates for each regression. For comparison, complete cases models are also reported in S.Table 5.

## Data Availability

All data produced in the present study are available from the Cleft Collective upon reasonable request.

## Acknowledgements

This research was funded in whole by the Medical Research Council MRC (MR/W020297/1). For the purpose of open access, the authors have applied a Creative Commons Attribution (CC BY) public copyright license to any Author Accepted Manuscript (AAM) version arising from this submission. RB, AR, SL and ES are employed in the MRC Integrative Epidemiology Unit (IEU) (MC_UU_00032/1 - Programme 1: Mendelian Randomization). SJL received support for this study from an MRC project grant (MR/T002093/1). CD is funded by the Wolfson Centre for Young People’s Mental Health, established with support from the Wolfson Foundation. MJO is funded by the Medical Research Council (MR/L010305/1 and programme grant MR/P005748/1). KK is funded by the Mental Health Mission. ER is funded by the UKRI MRC Programme Grant (MR/Y004094/1) and UKRI Future Leaders Fellowship Grant (MR/Y033922/1). MBMvdB is funded by the Medical Research Council (MR/V004905/1RE). MBMvdB also acknowledges funding from the Tackling Multimorbidity at Scale Strategic Priorities Fund programme (MR/W014416/1) delivered by the Medical Research Council and the National Institute for Health Research in partnership with the Economic and Social Research Council and in collaboration with the Engineering and Physical Sciences Research Council.

This publication involves data derived from independent research funded by The Scar Free Foundation; additional funding was provided by The Underwood Trust and the Vocational Training Charitable Trust (VTCT) (REC approval 13/SW/0064). We are grateful to the families who participated in the study, the UK NHS cleft teams, and The Cleft Collective team, who helped facilitate the study. The views expressed in this publication are those of the author(s) and not necessarily those of The Scar Free Foundation, The Underwood Trust, the Vocational Training Charitable Trust or The Cleft Collective Cohort Study team. For the data used from the Born in Bradford cohort study we want to acknowledge that Born in Bradford is only possible because of the enthusiasm and commitment of the children and parents in BiB. We are grateful to all the participants, health professionals, schools, and researchers who have made Born in Bradford happen.

This work was carried out using the computational facilities of the Advanced Computing Research Centre, University of Bristol - http://www.bristol.ac.uk/acrc/.

## Abbreviations

ALSPAC: Avon Longitudinal Study of Parents and Children
ASQ:SE-2: Ages and Stages Questionnaire - Social Emotional 2
ASQ-3: Ages and Stages Questionnaire 3
BiB: Born in Bradford
CI: Confidence Interval
CL/P: Cleft Lip and/or Palate
CNV: Copy Number Variant
GCSE: General Certificate of Secondary Education
GSA: Global Screening Array
kb: Kilobase
MCS: Millenium Cohort Study
ND CNV: Neurodevelopmental Copy Number Variant
QC: Quality Control
SDQ: Strengths and Difficulties questionnaire

