## Supplemental Tables and Figures for "Copy Number Variants and their Implications for Developmental and Behavioural Problems in Cleft Lip and/or Palate"

Supplementary Materials for “Copy Number Variants Linked to Neurodevelopmental Disorders and their Implications for Cleft Lip and/or Palate”.

Table of Contents

Note

Figures

Tables

References

### Supplementary Note 1: Neurodevelopmental CNV calling

We called CNVs using PennCNV 1.0.5 making use of biallelic markers common to Illumina and PennCNV. This procedure is outlined in S. Figure 2. To conduct CNV QC, we visualized all CNV calls and adjusted QC parameters to remove outliers from the distribution, see S. Figure 3. Specifically, samples were excluded based on 100 or more CNVs, waviness factor < -0.037 or >0.037, or log R ratio SD > 0.24. The value of the log R ratio standard deviation was defined based on two standard deviations from the mean of our log R ratio standard deviation to define a boundary of normally distributed high quality CNV calls. From 7182 raw samples, the final sample consisted of 6551, see S. Figure 1 and S. Figure 4. We plotted the B-allele Frequency and log R ratio of each sample with a potential neurodevelopmental CNV and visually inspected each plot to manually verify neurodevelopmental CNV status, see examples in S. Figure 5. We also visualized the plots of any samples which approximately met the criteria for any of the neurodevelopmental CNVs to ensure we did not miss samples with neurodevelopmental CNVs due to small margins around cut points.

### Supplemental Figure 1: CNV quality control flow diagram

52 failed PennCNV QC

**Raw samples 7182**

7126

7102

6918 samples into CNV call

56 controls

24 replicates

136 call rate < 0.97

6966

48 sex discordant

6866

**6551 samples with quality CNV calls**

367 fail CNV QC

N>100CNVs

LRR SD >0.24

Wavefactor < -0.03 or >0.03

### Supplemental Figure 2: Calling Neurodevelopmental CNV Steps

Step 1: Generate CNVs

Illumina GenomeStudio export LRR and B-Allele frequencies

Generate PNCV input files:

Sample Files

PFB

Gcmodel

Perform CNV call using detect_cnv.pl

Step 2: CNV Quality Control

Exclude individuals: N>100CNVs

LRR SD >0.24

Wavefactor < -0.04 or >0.04 Genotyping <0.95

Exclude CNVs: <20 probes Density per probe <20kb

Combine CNVs if the distance between CNVs is < 50% of their combined length

STEP 3: Call Neurodevelopmental CNVs

Cross-reference CNVs against coordinates for 54 neurodevelopmental CNVs

Identify whether these CNVs pass locus specific criteria(e.g. hitting critical genes, or >50% length of critical region)

Produce LRR/B-allele frequency trace plots for each ND CNV for visual inspection

*Genotyping and preliminary QC steps*

### Supplemental Figure 3: CNVs before and after CNV QC

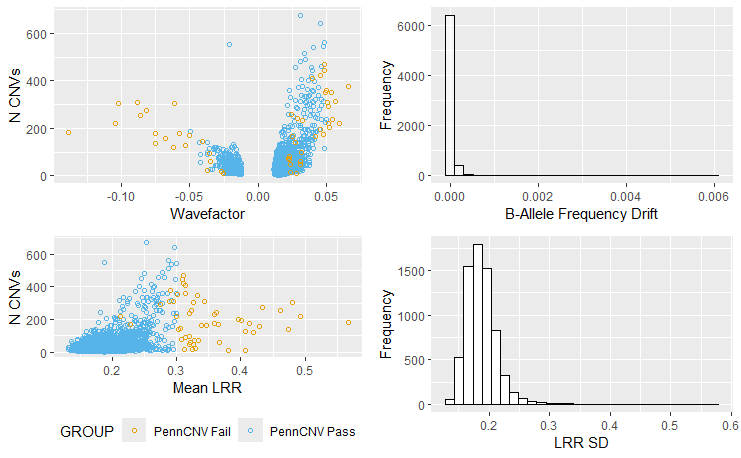

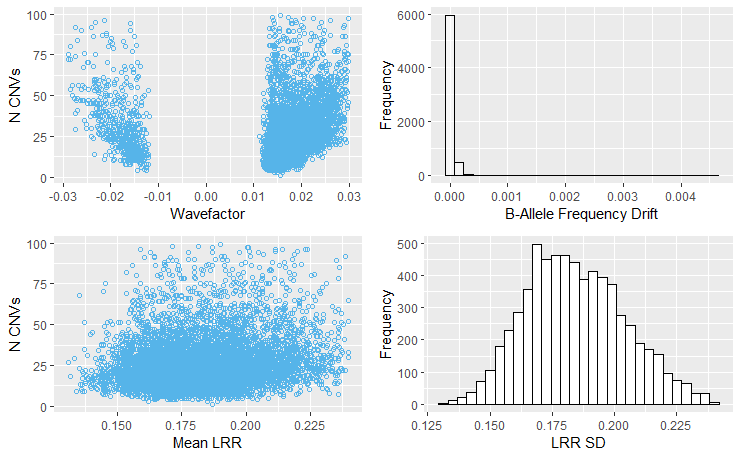

**After CNV QC, N = 6551**

**Before CNV QC, N = 6918**

### Supplemental Figure 4: Sample Exclusion Venn Diagram

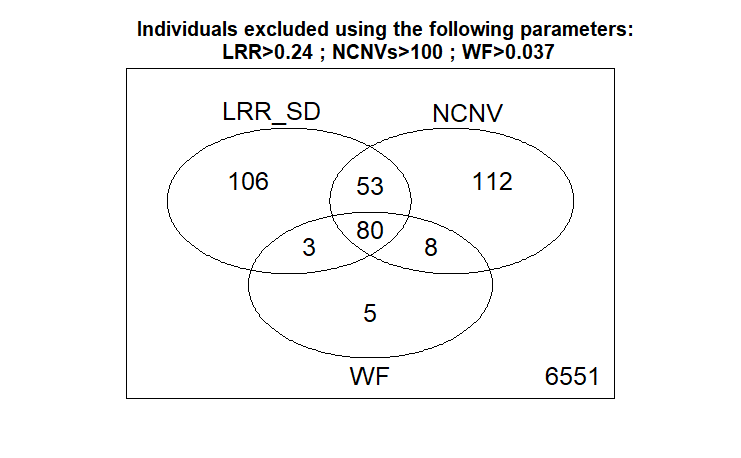

### Supplemental Figure 5: Examples of CNV probe plots for i) a 22q11 deletion and ii) a 22q11 duplication

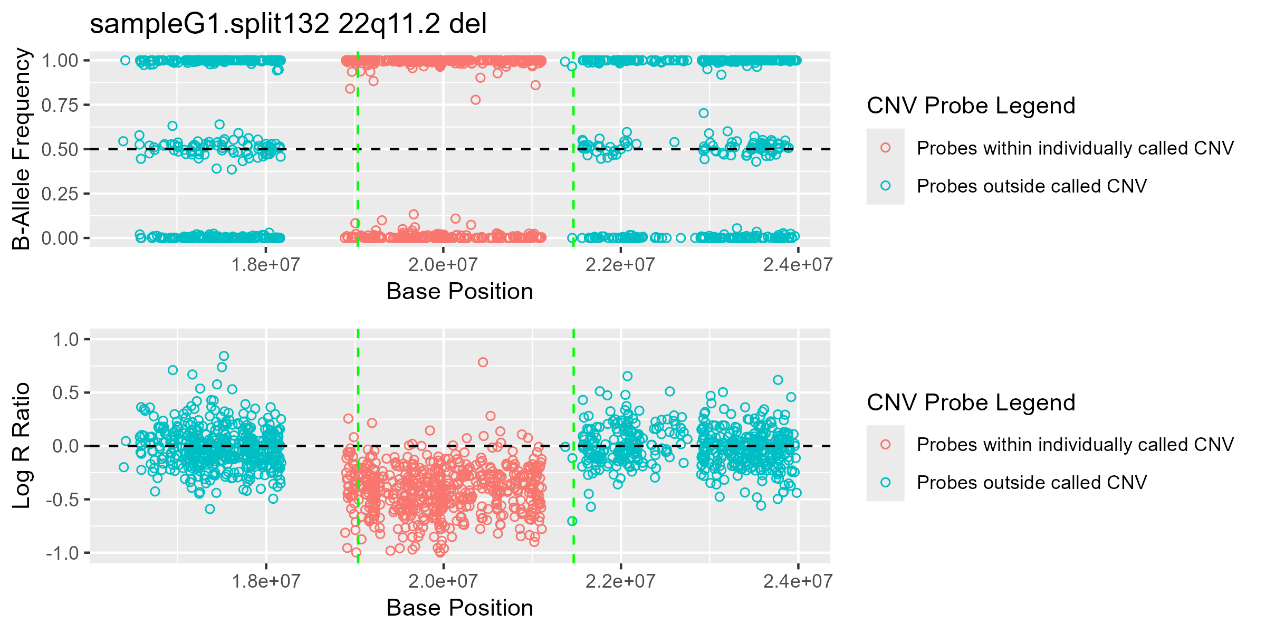

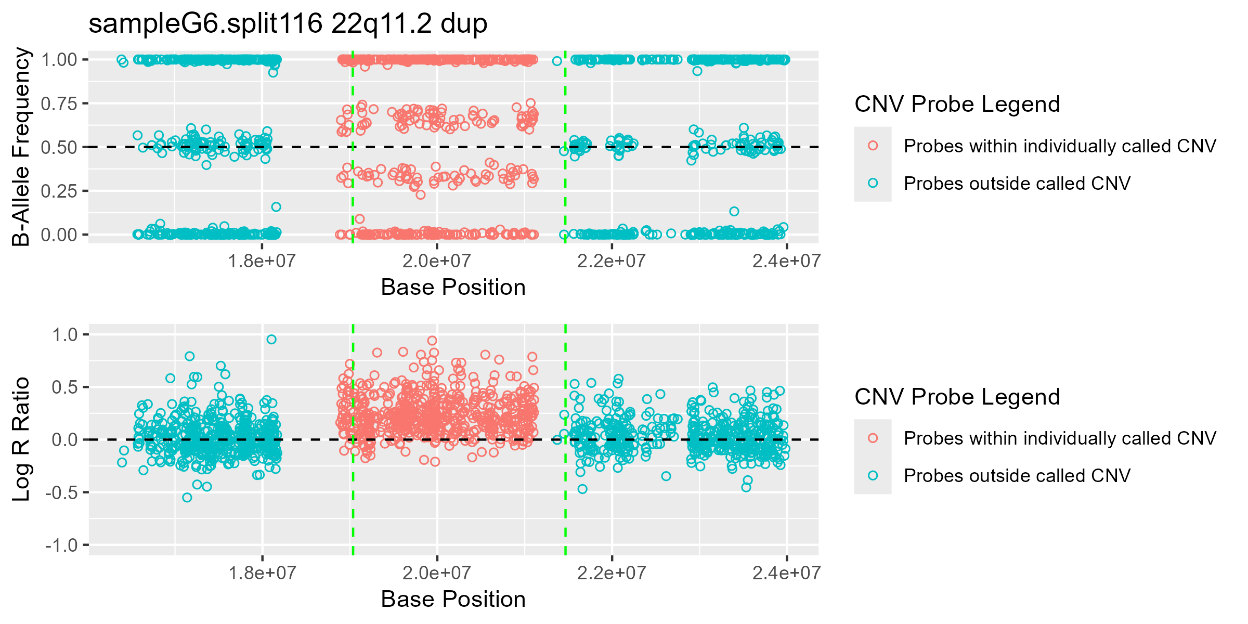

### Supplemental Figure 6: Pearson correlations between maternal (M) and paternal (P) ratings of Strengths and Difficulties Questionnaires (SDQ) and Ages and Stages: Social Emotional-2 (ASQ:SE - 2)

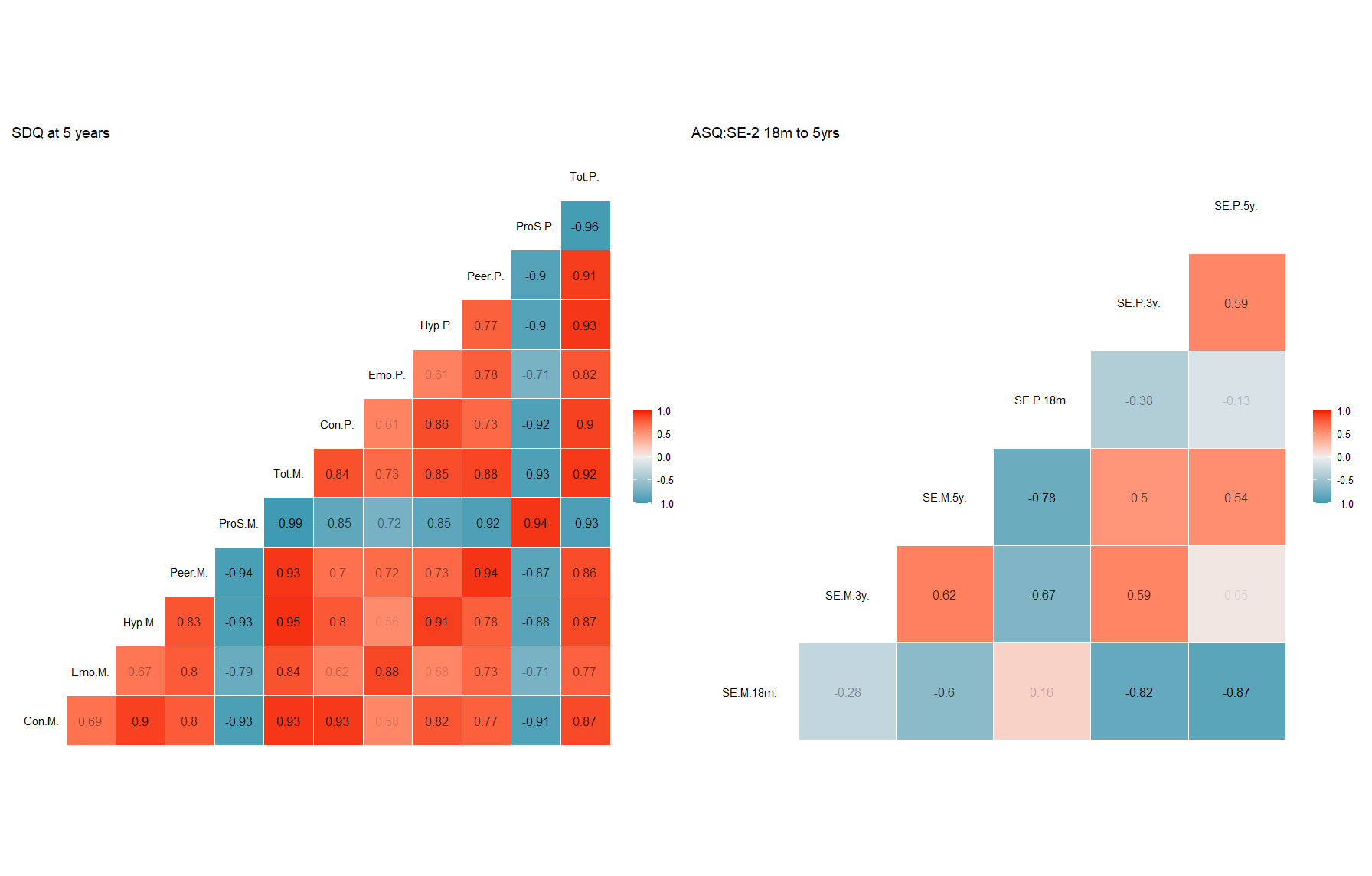

Subscale abbreviations are as follows; Tot = total score, ProS = Prosocial, Peer = Peer problems, Hyp= Hyperactivity, Emo = Emotional, Con= Conduct, SE = Social Emotional.

### Supplemental Figure 7: Pearson Correlations between maternal (M) and paternal (P) ratings Ages and Stages Questionnaire-3 (ASQ)

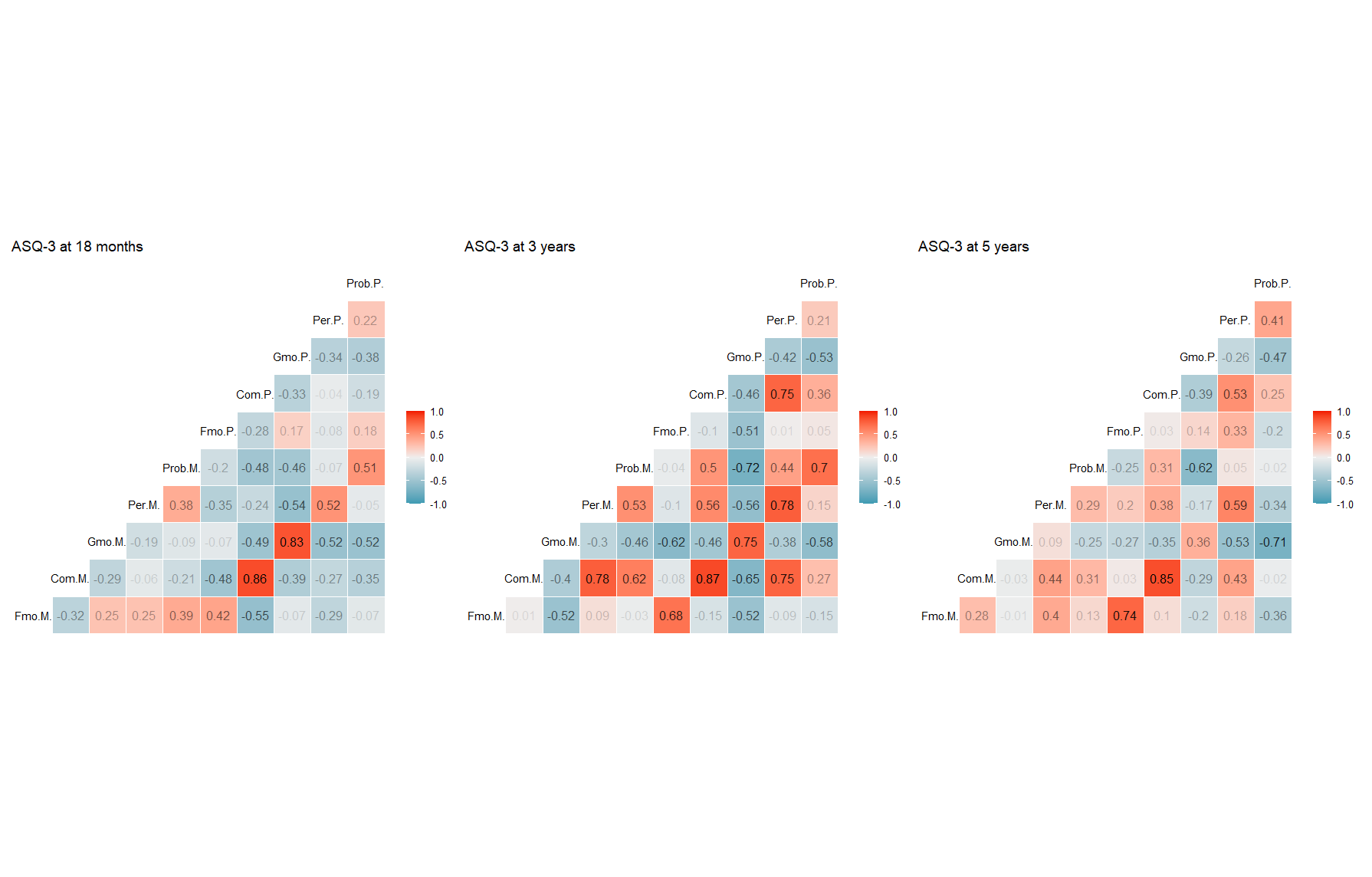

Subscale abbreviations are as follows; Prob = Problem Solving, Per = Personal Social, Gmo = Gross motor, Com= Communication, Fmo = Fine motor.

### Supplemental Table 1: Comparison of Cleft Collective individuals with and without genetic samples on key characteristics (1,729 individuals without genetics, 2,179 individuals with genetics, 3,908 in total)

| Categorical Measures | | | | |
| --- | --- | --- | --- | --- |
| Measure | N (Total) | Biggest Class in sample without genetics (%) | Biggest Class in sample with genetics (%) | x^2^ p-value |
| Cleft Type | 2713 | Cleft palate (40.1%) | Cleft palate (37.7%) | 0.41 |
| Syndromic | 2713 | No (91.7%) | No (91.7%) | 1 |
| Pierre Robin Sequence | 3313 | No (88.9%) | No (87.4%) | 0.29 |
| Maternal Ethnicity | 1332 | White (90%) | White (92%) | 0.5 |
| Maternal Highest Qual. (0-14) | 1336 | First Degree (30%) | First Degree (31%) | 0.7 |
| Measure | N(Total) | Median (Q1, Q3) | Median(Q1, Q3) | Wilcoxon rank sum p-value |
| Maternal Age at Birth | 1373 | 31 (27.0- 34.0) | 31 (27.0 - 34.0) | 0.3 |
| Measure | N (Total) | Mean (SD) | Mean (SD) | T-test p-value |
| ASQ- Social Emotional 2 (18m) | 1140 | 32.1 (26) | 29.8 (24) | 0.14 |
| ASQ- Social Emotional 2 (3y) | 1009 | 45.2(42.5) | 41.2 (42.1) | 0.2 |
| ASQ- Social Emotional 2 (5y) | 775 | 42.9 (51.4) | 39.5(43.9) | 0.41 |
| SDQ Total (5y) | 774 | 9.2 (6.3) | 9.01 (5.7) | 0.71 |
| SDQ Prosocial (5y) | 777 | 8 (2.2) | 7.75 (2.1) | 0.18 |
| Measure | N (Total) | Mean (SD) | Mean (SD) | T-test p-value |
| ASQ-3 Communication (18m) | 1185 | 30 (16.2) | 33.2 (15.9) | 9 x 10^-4^ |
| ASQ-3 Gross Motor (18m) | 1187 | 48.5 (17.3) | 51.5 (14.2) | 2 x 10^-3^ |
| ASQ-3 Fine Motor (18m) | 1183 | 59.5 (13.5) | 52.6 (11.5) | 7 x 10^-3^ |
| ASQ-3 Problem Solving (18m) | 1158 | 38.6 (16.2) | 41.1 (14.4) | 9 x 10^-3^ |
| ASQ-3 Personal-Social (18m) | 1183 | 44.3 (13) | 46.3 (11.1) | 5 x 10^-3^ |
| ASQ-3 Communication (3yr) | 1019 | 49.8 (14) | 50.2 (13.3) | 0.64 |
| ASQ-3 Gross Motor (3yr) | 1016 | 52.1 (14.5) | 53.3 (11.6) | 0.20 |
| ASQ-3 Fine Motor (3yr) | 1008 | 43.3 (17.5) | 44.6 (15.9) | 0.27 |
| ASQ-3 Problem Solving (3yr) | 1012 | 51.5 (14) | 53 (12.3) | 0.11 |
| ASQ-3 Personal-Social (3yr) | 1018 | 50 (12.7) | 51.1(11.3) | 0.21 |
| ASQ-3 Communication (5yr) | 780 | 52.8 (13.6) | 53.9 (13.1) | 0.33 |
| ASQ-3 Gross Motor (5yr) | 779 | 50.3 (13.4) | 49.5 (14.4) | 0.55 |
| ASQ-3 Fine Motor (5yr) | 783 | 49.8 (15.2) | 50.5 (15.6) | 0.57 |
| ASQ-3 Problem Solving (5yr) | 781 | 53.7 (11.6) | 53.8 (12) | 0.91 |
| ASQ-3 Personal-Social (5yr) | 784 | 52.3 (11.9) | 53.1 (11.4) | 0.41 |

### Supplemental Table 2: Neurodevelopmental CNVs from (Kendall, Rees et al. 2019)

| **Neurodevelopmental CNV Reference List** | | |
| --- | --- | --- |
| 1p36 del (GABRD) | 1p36 | chr1:0-2500000 |
| 1p36 dup (GABRD) | 1p36 | chr1:0-2500000 |
| TAR del | 1q21.1 | chr1:145394955-145807817 |
| TAR dup | 1q21.1 | chr1:145394955-145807817 |
| 1q21.1 del | 1q21.1 | chr1:146527987-147394444 |
| 1q21.1 dup | 1q21.1 | chr1:146527987-147394444 |
| NRXN1 del | 2p16.3 | chr2:50145643-51259674 |
| 2q11.2 del (LMAN2L, ARID5A) | 2q11.2 | chr2:96742409-97677516 |
| 2q13 del | 2q13 | chr2:111394040-112012649 |
| 2q13 dup | 2q13 | chr2:111394040-112012649 |
| 2q37 del (HDAC4) | 2q37 | chr2:239716679-243199373 |
| 3q29 del | 3q29 | chr3:195720167-197354826 |
| Wolf_Hirschhorn del | 4p16.3 | chr4:1552030-2091303 |
| Wolf_Hirschhorn dup | 4p16.3 | chr4:1552030-2091303 |
| Sotos syndrome del | 5q35 | chr5:175720924-177052594 |
| WBS del | 7q11.23 | chr7:72744915-74142892 |
| WBS dup | 7q11.23 | chr7:72744915-74142892 |
| 8p23.1 del | 8p23.1 | chr8:8098990-11872558 |
| 8p23.1 dup | 8p23.1 | chr8:8098990-11872558 |
| 9q34 del (EHMT1) | 9q34 | chr9:140513444-140730578 |
| 10q23 del (NRG3, GRID1) | 10q22q23 | chr10:82045472-88931651 |
| Potocki-Shaffer syndrome del (EXT2) | 11p11.2 | chr11:43940000-46020000 |
| 15q11.2 del BP1-BP2 | 15q11.2 | chr15:22805313-23094530 |
| 15q11.2 dup BP1-BP2 | 15q11.2 | chr15:22805313-23094530 |
| PWS_AS del | 15q11.2q12 | chr15:22805313-28390339 |
| PWS_AS dup | 15q11.2q12 | chr15:22805313-28390339 |
| 15q13.3 del BP4-BP5 | 15q13.3 | chr15:31080645-32462776 |
| 15q24 del | 15q24 | chr15:72900171-78151253 |
| 15q24 dup | 15q24 | chr15:72900171-78151253 |
| 15q25 del | 15q25.2 | chr15:83219735-85722039 |
| 16p13.11 del | 16p13.11 | chr16:15511655-16293689 |
| 16p13.11 dup | 16p13.11 | chr16:15511655-16293689 |
| 16p12.1 del | 16p12.1 | chr16:21950135-22431889 |
| 16p11.2 distal del | 16p11.2 | chr16:28823196-29046783 |
| 16p11.2 distal dup | 16p11.2 | chr16:28823196-29046783 |
| 16p11.2 del | 16p11.2 | chr16:29650840-30200773 |
| 16p11.2 dup | 16p11.2 | chr16:29650840-30200773 |
| 17p13.3 del (YWHAE) | 17p13.3 | chr17:1247834-1303556 |
| 17p13.3 dup (YWHAE) | 17p13.3 | chr17:1247834-1303556 |
| 17p13.3 del (PAFAH1B1) | 17p13.3 | chr17:2496923-2588909 |
| 17p13.3 dup (PAFAH1B1) | 17p13.3 | chr17:2496923-2588909 |
| Potocki-Lupski syndrome dup | 17p11.2 | chr17:16812771-20211017 |
| Smith-Magenis syndrome del | 17p11.2 | chr17:16812771-20211017 |
| 17q11.2 del (NF1) | 17q11.2 | chr17:29107491-30265075 |
| 17q11.2 dup (NF1) | 17q11.2 | chr17:29107491-30265075 |
| Renal cysts and diabetes syndrome del | 17q12 | chr17:34815904-36217432 |
| 17q12 dup | 17q12 | chr17:34815904-36217432 |
| 17q21.31 del | 17q21.31 | chr17:43705356-44164691 |
| 22q11.2 del | 22q11.2 | chr22:19037332-21466726 |
| 22q11.2 dup | 22q11.2 | chr22:19037332-21466726 |
| 22q11.2 distal del | 22q11.2 | chr22:21920127-23653646 |
| 22q11.2 distal dup | 22q11.2 | chr22:21920127-23653646 |
| SHANK3 del | 22q13 | chr22:51113070-51171640 |
| SHANK3 dup | 22q13 | chr22:51113070-51171640 |

### Supplemental Table 3: Fit statistics for multilevel models of Ages and Stages Questionnaire -3 (ASQ-3) and Ages and Stages Questionnaire - Social Emotional -2 (ASQ:SE-2)

*Starting with a null model for each subscale we incrementally included each term in the first column testing how each term impacted the fit of the data to the model using Akaike Information Criterion (AIC) and Loglikelihood Ratio Tests (LRT). Where a term improved model fit by reducing AIC and showing an LRT p-value below <0.05 we included the term in the final model as shown in the model formula.*

|  | **AIC** | **LRT p-value** | **Final model formula** |
| --- | --- | --- | --- |
| **ASQ-3 Communication** |  |  | communication ~ 1 + age + sex +sex*age + NDD_CNV + cleft_subtype + (1 \| ppt) + (1 \| occasion) |
| Null Model | 16339 |  |  |
| *Random Intercep*t | 15649 | <0.001 |  |
| Random Slope | 15653 | 1.000 |  |
| *Sex* | 15631 | <0.001 |  |
| *Sex*age* | 15626 | 0.005 |  |
| neurodevelopmental *CNV* | 15617 | 0.002 |  |
| *Cleft sub-type* | 15566 | <0.001 |  |
| **ASQ-3 Fine Motor** |  |  | fine_motor ~ 1 + age + sex + sex*age + NDD_CNV + cleft_subtype + (1 \| ppt) + (1 \| occasion) |
| Null Model | 15625 |  |  |
| *Random Intercept* | 15618 | 0.003 |  |
| Random Slope | 15862 | 1.000 |  |
| *Sex* | 15601 | <0.001 |  |
| *Sex*age* | 15590 | <0.001 |  |
| neurodevelopmental *CNV* | 15581 | <0.001 |  |
| *Cleft sub-type* | 15536 | <0.001 |  |
| **ASQ-3 Gross Motor** |  |  | gross_motor ~ 1 + age + sex + sex*age + NDD_CNV + cleft_subtype + (1 + age \| ppt) + (1 \| occasion) |
| Null Model | 15129 |  |  |
| *Random Intercept* | 15128 | 0.070 |  |
| *Random Slope* | 15099 | <0.001 |  |
| *Sex* | 15099 | <0.001 |  |
| *Sex*age* | 15094 | 0.006 |  |
| neurodevelopmental *CNV* | 15090 | 0.012 |  |
| *Cleft sub-type* | 15077 | <0.001 |  |
| **ASQ-3 Personal Social** |  |  | personal_social ~ 1 + age + sex + sex*age + NDD_CNV + cleft_subtype + (1 + age \| ppt) + (1 \| occasion) |
| Null Model | 14663 |  |  |
| *Random Intercept* | 14488 | <0.001 |  |
| *Random Slope* | 14482 | 0.008 |  |
| *Sex* | 14472 | <0.001 |  |
| *Sex*age* | 14470 | 0.049 |  |
| neurodevelopmental *CNV* | 14467 | 0.002 |  |
| *Cleft sub-type* | 14430 | <0.001 |  |
| **ASQ-3 Problem Solving** |  |  | problem_solving ~ 1 + age + sex + sex*age + NDD_CNV + cleft_subtype + (1 \| ppt) + (1 \| occasion) |
| Null Model | 15805 |  |  |
| *Random Intercept* | 15554 | <0.001 |  |
| Random Slope | 15558 | 1.000 |  |
| *Sex* | 15551 | 0.032 |  |
| *Sex*age* | 15549 | 0.038 |  |
| neurodevelopmental *CNV* | 15543 | 0.004 |  |
| *Cleft sub-type* | 15502 | <0.001 |  |
| **ASQ: SE-2** |  |  | asq_se ~ 1 + age + sex + NDD_CNV + cleft_subtype + (1 + age \| ppt) + (1 \| occasion) |
| *Null Model* | 19081 |  |  |
| *Random Intercept* | 19033 | <0.001 |  |
| *Random Slope* | 18744 | <0.001 |  |
| *Sex* | 18729 | <0.001 |  |
| *Sex*age* | 18719 | 0.162 |  |
| neurodevelopmental *CNV* | 18710 | 0.010 |  |
| *Cleft sub-type* | 18665 | <0.001 |  |

### Supplemental Table 4: Fixed effect estimates of cleft sub-type in linear multilevel models of the Ages and Stages Questionnaire -3 (ASQ-3) and Ages and Stages Questionnaire - Social Emotional -2 (ASQ:SE-2).

The reference category was cleft lip only. Sex, age and neurodevelopmental copy number variants were included as covariates. Higher scores on ASQ-3 domains indicate better developmental outcomes while higher scores on ASQ:SE-2 indicate higher risk of impairment in early social and emotional development.

| *Development 18m to 5 years* | Coefficient | 95%CI | p-value |
| --- | --- | --- | --- |
| ASQ-3 Communication | | | |
| Cleft palate | -5.44 | -7.64, -3.24 | 1.26x10^-6^ |
| Cleft lip and palate | -3.90 | -6.06, -1.74 | 4x10^-4^ |
| Submucous cleft palate | -11.66 | -20.88, -2.44 | 0.013 |
| ASQ-3 Fine Motor | | | |
| Cleft palate | -4.12 | -6.24, -1.99 | 1.4x10^-4^ |
| Cleft lip and palate | -2.55 | -4.63, -0.46 | 0.017 |
| Submucous cleft palate | -6.94 | -15.89, 2.02 | 0.129 |
| ASQ-3 Gross Motor | | | |
| Cleft palate | -3.96 | -6.04, -1.89 | 1.8x10^-4^ |
| Cleft lip and palate | -1.07 | -3.10, 0.97 | 0.304 |
| Submucous cleft palate | -9.25 | -17.86, -0.65 | 0.035 |
| ASQ-3 Personal Social | | | |
| Cleft palate | -3.72 | -5.78, -1.67 | 3.9x10^-4^ |
| Cleft lip and palate | -2.06 | -4.07, -0.05 | 0.045 |
| Submucous cleft palate | -9.94 | -18.61, -1.26 | 0.025 |
| ASQ-3 Problem Solving | | | |
| Cleft palate | -2.69 | -4.46, -0.92 | 0.003 |
| Cleft lip and palate | -1.06 | -2.79, 0.67 | 0.230 |
| Submucous cleft palate | -2.13 | -9.39, 5.13 | 0.565 |
| ASQ: Social Emotional-2 | | | |
| Cleft palate | 6.41 | 1.37, 11.45 | 0.013 |
| Cleft lip and palate | 3.77 | -1.13, 8.68 | 0.132 |
| Submucous cleft palate | 14.31 | -5.38, 34.01 | 0.154 |

### Supplemental Table 5: Neurodevelopmental CNV and Strengths and Difficulties regression models in the complete (non-imputed) dataset.

| *SDQ at 5 years* | ND CNV | No ND CNV | Estimate | 95% Confidence Interval | P-value |
| --- | --- | --- | --- | --- | --- |
| Conduct Problems | 21 | 562 | 1.17 | 0.38, 1.95 | 0.004 |
| Emotional | 21 | 562 | 0.98 | 0.10, 1.86 | 0.029 |
| Hyperactivity | 20 | 561 | 0.99 | -0.27, 2.26 | 0.122 |
| Peer Problems | 21 | 563 | 1.69 | 0.84, 2.54 | 1.1x10^-4^ |
| Prosocial | 21 | 562 | -2.21 | -3.20, -1.22 | 1.4x10^-5^ |
| SDQ Total | 20 | 561 | 5.27 | 2.31, 8.22 | 5x10^-4^ |

Estimated effect of having a neurodevelopmental copy number variant (ND CNV) in linear regression models of the Strengths and Difficulties Questionnaire (SDQ) on complete cases data at age 5. The reference category is no ND CNV. Please note higher scores on the SDQ indicate higher risk of behavioural problems except for prosocial subscale where higher scores indicate fewer prosocial traits.

### Supplemental Table 6: Complete case linear regression analysis of age 5 Strengths and Difficulties Questionnaire scores by cleft type

The reference category is cleft lip only. Sex, age and neurodevelopmental copy number variants were included as covariates. Higher scores on SDQ subscales indicate higher risk of behavioral problems with the exception of the prosocial subscale, where higher scores indicate lower risk of behavioral problems.

|  | Estimate | 95%CI | p-value |
| --- | --- | --- | --- |
| Conduct Problems (N = 21 ND CNV/ 562 No ND CNV) | | | |
| Cleft palate | 0.17 | -0.22, 0.56 | 0.387 |
| Cleft lip and palate | 0.36 | -0.04, 0.75 | 0.074 |
| Submucous cleft palate | 0.05 | -1.70, 1.81 | 0.951 |
| Emotional (N = 21 ND CNV/562 No ND CNV) | | | |
| Cleft palate | 0.35 | -0.09, 0.78 | 0.118 |
| Cleft lip and palate | 0.13 | -0.30, 0.57 | 0.548 |
| Submucous cleft palate | 1.23 | -0.73, 3.19 | 0.217 |
| Hyperactivity (N = 20 ND CNV/561 No ND CNV) | | | |
| Cleft palate | 0.62 | 0.01, 1.23 | 0.045 |
| Cleft lip and palate | 0.65 | 0.04, 1.26 | 0.038 |
| Submucous cleft palate | 1.19 | -1.56, 3.93 | 0.395 |
| Peer Problems (N = 21 ND CNV/563 No ND CNV) | | | |
| Cleft palate | 0.39 | -0.03, 0.81 | 0.068 |
| Cleft lip and palate | 0.34 | -0.08, 0.76 | 0.114 |
| Submucous cleft palate | 0.39 | -1.50, 2.29 | 0.684 |
| Peer Problems (N = 21 ND CNV/562 No ND CNV) | | | |
| Cleft palate | -0.60 | -1.08, -0.11 | 0.016 |
| Cleft lip and palate | -0.60 | (-1.08, -0.11) | 0.016 |
| Submucous cleft palate | -1.44 | (-3.63, 0.75) | 0.197 |
| SDQ Total (N = 20 ND CNV/561 No ND CNV) | | | |
| Cleft palate | 1.51 | (0.08, 2.93) | 0.038 |
| Cleft lip and palate | 1.51 | (0.08, 2.93) | 0.038 |
| Submucous cleft palate | 2.89 | (-3.53, 9.32) | 0.377 |

### Supplemental Table 7: Frequency of all neurodevelopmental CNVs found by Cohort

| **Locus** | **Cleft Collective  (N = 2,180)** | **ALSPAC  (N = 6,361)** | **BiB (N = 7,626)** | **MCS (N = 6,710)** | **UK Biobank  (N = 151,619)** |
| --- | --- | --- | --- | --- | --- |
| 10q23 del (NRG3, GRID1) | 0 | 0 | 0 | 0 | 0 |
| 15q11.2 del BP1-BP2 | 17 | 43 | 29 | 52 | 544 |
| 15q11.2 dup BP1-BP2 | 13 | 32 | 44 | 38 | 762 |
| 15q13.3 del BP4-BP5 | <5 | <5 | <5 | <5 | 91 |
| 15q24 del | 0 | 0 | 0 | 0 | <5 |
| 15q24 dup | 0 | <5 | 0 | 0 | <5 |
| 15q25 del | 0 | 0 | 0 | 0 | 0 |
| 16p11.2 del | 10 | <5 | <5 | <5 | 43 |
| 16p11.2 distal del | <5 | <5 | <5 | 0 | 18 |
| 16p11.2 distal dup | <5 | 0 | <5 | 5 | 45 |
| 16p11.2 dup | <5 | <5 | 0 | 5 | 43 |
| 16p12.1 del | <5 | 5 | 8 | 5 | 86 |
| 16p13.11 del | <5 | <5 | 7 | <5 | 52 |
| 16p13.11 dup | <5 | 15 | 13 | 18 | 303 |
| 17p13.3 del (PAFAH1B1) | 0 | 0 | 0 | 0 | <5 |
| 17p13.3 del (YWHAE) | 0 | 0 | 0 | 0 | 17 |
| 17p13.3 dup (PAFAH1B1) | 0 | 0 | 0 | 0 | <5 |
| 17p13.3 dup (YWHAE) | 0 | 0 | 0 | 0 | 7 |
| 17q11.2 del (NF1) | 0 | 0 | 0 | 0 | <5 |
| 17q11.2 dup (NF1) | 0 | 0 | 0 | 0 | <5 |
| 17q12 dup | 0 | <5 | 0 | 0 | 35 |
| 17q21.31 del | 0 | 0 | 0 | 0 | 8 |
| 1p36 del (GABRD) | 0 | <5 | <5 | 0 | 0 |
| 1p36 dup (GABRD) | 0 | 0 | 0 | 0 | 0 |
| 1q21.1 del | <5 | 7 | <5 | <5 | 41 |
| 1q21.1 dup | <5 | 5 | 9 | 5 | 67 |
| 22q11.2 del | 10 | 0 | <5 | <5 | 5 |
| 22q11.2 distal del | <5 | 0 | 0 | 0 | <5 |
| 22q11.2 distal dup | 0 | 0 | 0 | <5 | <5 |
| 22q11.2 dup | 6 | 7 | 7 | 6 | 97 |
| 2q11.2 del (LMAN2L, ARID5A) | 0 | 0 | 0 | 0 | 11 |
| 2q13 del | 0 | <5 | <5 | <5 | 16 |
| 2q13 dup | <5 | <5 | <5 | <5 | 29 |
| 2q37 del (HDAC4) | 0 | 0 | 0 | 0 | 0 |
| 3q29 del | 0 | 0 | 0 | 0 | 5 |
| 8p23.1 del | 0 | 0 | <5 | 0 | 0 |
| 8p23.1 dup | <5 | 0 | <5 | 0 | <5 |
| 9q34 del (EHMT1) | 0 | 0 | 0 | 0 | 0 |
| NRXN1 del 2p16.3 | <5 | <5 | <5 | 5 | 53 |
| Potocki-Lupski syndrome dup (17p11.2) | 0 | 0 | 0 | 0 | <5 |
| Potocki-Shaffer syndrome del (EXT2) (11p11.2) | 0 | 0 | 0 | 0 | 0 |
| Prader-Willi syndrome/Angelman syndrome del (15q11.2q12) | 0 | 0 | 0 | 0 | 0 |
| Prader-Willi syndrome/Angelman syndrome dup (15q11.2q12) | 0 | 0 | 0 | 0 | 9 |
| Renal cysts and diabetes syndrome del (17q12) | 0 | 0 | <5 | 0 | <5 |
| SHANK3 del (22q13) | 0 | 0 | 0 | 0 | 0 |
| SHANK3 dup (22q13) | 0 | 0 | 0 | 0 | 0 |
| Smith-Magenis syndrome del (17p11.2) | 0 | 0 | 0 | 0 | 0 |
| Sotos syndrome del (5q35) | 0 | 0 | 0 | 0 | 0 |
| TAR del 1q21.2 | 0 | <5 | <5 | 0 | 27 |
| TAR dup 1q21.1 | 0 | 5 | <5 | 0 | 139 |
| Williams-Beuren syndrome del (7q11.23) | 0 | 0 | 0 | 0 | 5 |
| Williams-Beuren syndrome dup (7q11.23) | 0 | 0 | 0 | 0 | <5 |
| Wolf-Hirschhorn del (4p16.3) | 0 | 0 | 0 | 0 | 0 |
| Wolf-Hirschhorn dup (4p16.3) | 0 | 0 | 0 | 0 | <5 |

Abbreviations: CNV; copy number variant, ALSPAC; Avon Longitudinal Study of Parents and Children, BiB; Born in Bradford, MCS; Millenium Cohort Study, del; deletion, dup; duplication, NRG3; Neuregulin 3, GRID1; Glutamate Ionotropic Receptor Delta Type Subunit 1, PAFAH1B1; platelet activating factor acetylhydrolase 1b regulatory subunit 1, YWHAE; Tyrosine 3-Monooxygenase/Tryptophan 5-Monooxygenase Activation Protein Epsilon, NF1; Neurofibromin 1, GABRD; Gamma-Aminobutyric Acid Type A Receptor Subunit Delta, LMAN2L; Lectin, Mannose Binding 2 Like, ARID5A; AT-Rich Interaction Domain 5A, HDAC4; Histone Deacetylase 4, EHMT1; Euchromatic Histone Lysine Methyltransferase 1, NRXN1; Neurexin 1, EXT2; Exostosin Glycosyltransferase 2, SHANK3; SH3 And Multiple Ankyrin Repeat Domains 3, TAR; thrombocytopenia with absent radius.

### Supplemental Table 8: Odds ratios of having a neurodevelopmental copy number variant (ND CNV) among parents and siblings from the Cleft Collective compared to general population comparison groups

| Cohort |  | ND CNV | No ND CNV | Percentage of included with ND CNVs | Odds Ratio | 95 % Confidence intervals | P-value |
| --- | --- | --- | --- | --- | --- | --- | --- |
| **Cleft Collective Mothers** |  | 54 | 2,290 | 2.3% | reference | reference | reference |
| ALSPAC |  | 144 | 6,217 | 2.3% | 1.00 | 0.72, 1.39 | 0.872 |
| BiB |  | 149 | 7,477 | 2.0% | 1.18 | 0.85, 1.63 | 0.315 |
| MCS |  | 151 | 6,559 | 2.3% | 1.02 | 0.73, 1.14 | 0.872 |
| UK Biobank |  | 2,583 | 149,036 | 1.7% | 1.38 | 1.03, 1.82 | 0.023 |
| **Cleft Collective Fathers** |  | 54 | 1,815 | 3.0% | reference | reference | reference |
| ALSPAC |  | 144 | 6,217 | 2.3% | 1.27 | 0.90, 1.75 | 0.146 |
| BiB |  | 149 | 7,477 | 2.0% | 1.49 | 1.07, 2.06 | 0.016 |
| MCS |  | 151 | 6,559 | 2.3% | 1.29 | 0.92, 1.78 | 0.122 |
| UK Biobank |  | 2,583 | 149,036 | 1.7% | 1.75 | 1.30, 2.30 | 1.2x10^-4^ |
| **Cleft Collective Siblings** |  | 5 | 153 | 3.4% | reference | reference | reference |
| ALSPAC |  | 144 | 6,217 | 2.3% | 1.48 | 0.47, 3.61 | 0.398 |
| BiB |  | 149 | 7,477 | 2.0% | 1.74 | 0.55, 4.25 | 0.222 |
| MCS |  | 151 | 6,559 | 2.3% | 1.51 | 0.48, 3.68 | 0.393 |
| UK Biobank |  | 2,583 | 149,036 | 1.7% | 2.04 | 0.65, 4.87 | 0.107 |

Abbreviations: ND CNV; Neurodevelopmental copy number variant, ALSPAC; Avon Longitudinal Study of Parents and Children, BiB; Born in Bradford, MCS; Millenium Cohort Study

### Supplemental Table 9: Fixed effect estimates of inherited vs de novo neurodevelopmental copy number variants (ND CNVs) in linear multilevel models of Ages and Stages Questionnaire -3 (ASQ-3) and Ages and Stages Questionnaire - Social Emotional -2 (ASQ:SE-2).

The reference category is no ND CNV. Please note higher scores on the ASQ-3 domains indicate better developmental outcomes and higher scores on ASQ:SE-2 indicate poorer social emotional outcomes. (N = 31 Inherited, 19 *De Novo,* 29 of Unknown Origin)

| Development 18m to 5 years | Coefficient | 95% CIs | p-value |
| --- | --- | --- | --- |
| ASQ-3 Communication | | | |
| *De novo* | -3.89 | -11.98, 4.20 | 0.346 |
| Inherited | -2.14 | -8.89, 4.60 | 0.533 |
| Unknown | -13.91 | -21.27, -6.55 | 2.1x10^-4^ |
| ASQ-3 Fine Motor | | | |
| *De novo* | 0.46 | -7.50, 8.41 | 0.910 |
| Inherited | -6.40 | -12.91, 0.12 | 0.054 |
| Unknown | -13.04 | -20.15, -5.92 | 3.3x10^-4^ |
| ASQ-3 Gross Motor | | | |
| *De novo* | -7.13 | -14.79, 0.53 | 0.068 |
| Inherited | 0.35 | -6.00, 6.70 | 0.914 |
| Unknown | -8.78 | -15.69, -1.87 | 0.013 |
| ASQ-3 Personal Social | | | |
| *De novo* | -1.78 | -9.36, 5.80 | 0.645 |
| Inherited | -4.67 | -10.97, 1.64 | 0.147 |
| Unknown | -9.74 | -16.64, -2.85 | 0.006 |
| ASQ-3 Problem Solving | | | |
| *De novo* | -1.46 | -8.01, 5.10 | 0.663 |
| Inherited | -3.71 | -9.14, 1.72 | 0.181 |
| Unknown | -6.71 | -12.61, -0.80 | 0.026 |
| ASQ: Social Emotional-2 | | | |
| *De novo* | 7.51 | -15.30, 30.33 | 0.519 |
| Inherited | 18.22 | -0.66, 37.10 | 0.059 |
| Unknown | 45.77 | 24.55, 66.99 | 2.36 x10^-5^ |

### Supplemental Table 10: Ages and Stages Questionnaire -3 (ASQ-3) and Ages and Stages Questionnaire - Social Emotional -2 (ASQ:SE-2) thresholds for monitoring and referral for developmental delays and social emotional problems

|  |  | 18 months | 3 years | 5 years |
| --- | --- | --- | --- | --- |
| ASQ-3 Communication | Monitoring | 13.06 | 30.99 | 33.19 |
| ASQ-3 Gross Motor | Monitoring | 37.38 | 36.99 | 31.28 |
| ASQ-3 Fine Motor | Monitoring | 34.32 | 18.07 | 26.54 |
| ASQ-3 Problem Solving | Monitoring | 25.74 | 30.29 | 29.99 |
| ASQ-3 Personal-Social | Monitoring | 27.19 | 35.33 | 39.07 |
| ASQ: Social Emotional - 2 | Monitoring | 50 | 75 | 70 |
|  | Referral | 65 | 105 | 95 |
